# Prospective genomic and epidemiologic surveillance of *Klebsiella pneumoniae* in a tertiary NICU

**DOI:** 10.64898/2025.12.10.25336365

**Authors:** Carolin Böhne, Claas Baier, Jelena Erdmann, Ella Ebadi, Carina Zirkler, Marc Lindenberg, Dirk Schlüter, Sabine Pirr, Corinna Peter, Bettina Bohnhorst, Leonard Knegendorf

## Abstract

**Background:** *Klebsiella pneumoniae* complex (Kp) is a relevant neonatal pathogen colonizing preterm infants. While outbreak investigations often focus on multidrug-resistant strains, the epidemiology and genomic dynamics of wild-type Kp in nonoutbreak neonatal intensive care unit (NICU) settings remain elusive.

**Methods:** We conducted a 30-month (October 2021 to March 2024) cohort study with weekly active, unselective colonization surveillance of all NICU patients to identify risk factors for nosocomial Kp acquisition and drivers of transmission in a tertiary 21-bed NICU/intermediate care unit (IMC) in Germany.

**Results:** Among 936 patients, 8.7% carried Kp, of which 70.4% were nosocomial. Very low birth weight (VLBW; <1500 g) was the only independent risk factor for nosocomial acquisition (adjusted odds ratio [aOR], 3.42; 95% CI, 1.29-9.32). Kp infections occurred in three Kp carriers (3.7%). Genomic analyses of at least the first isolate per patient (83 in total) revealed an oligoclonal population structure, with distinct sequence types driving temporally overlapping clusters. Ten genomic clusters (median size, four patients) were identified, with markedly higher odds in VLBW infants (aOR, 8.76; 95% CI, 2.45-34.16). Nosocomial cluster-assigned cases had higher rates and longer durations of noninvasive ventilation and peripheral venous catheter use. Cluster prevalence showed climate-associated variation, with a six-feature extreme gradient boosting (XGBoost) model identifying temperature and humidity among the strongest predictors.

**Conclusions:** Patient- and climate-associated parameters are main drivers of nosocomial wild-type Kp acquisition and cluster occurrence. Comprehensive surveillance and risk-adapted infection prevention and control support sustainable Kp control in VLBW infants.

## Background

The Gram-negative *Klebsiella pneumoniae* complex (Kp) is known for colonizing preterm infants in neonatal intensive care units (NICUs) (1) and is one of the pathogens frequently leading to neonatal infections (2, 3). Besides patient-related factors and the use of invasive devices and procedures (4–6), infection susceptibility is further influenced by pathogen-related attributes such as hypervirulent Kp isolates (hvKp) (7). A gene-based grading system (Kleborate) (8) and a hypermucoviscous Kp phenotype (positive string test (9) or tellurite resistance (10)), may indicate increased virulence. Hypervirulence may also coincide with antibiotic resistance, including Carbapenem resistance (11).

Kp can persist in the environment (e.g. surfaces, medical products), thus being easily transmittable in NICUs (12, 13) and resulting in several outbreak reports from NICUs worldwide (13–16), including Carbapenem-resistant Kp (17, 18). Since neonatal infections still represent a major cause of neonatal morbidity and mortality even in western, industrialized countries (19), close epidemiologic and molecular Kp monitoring in NICUs seems crucial from a clinical and infection prevention and control (IPC) perspective. In Germany, weekly microbiological colonization screening is mandatory in NICUs, following national recommendations of the Commission for Infection Prevention and Hygiene in Healthcare and Nursing (KRINKO) (20, 21). To clarify transmission pathways, whole-genome sequencing (WGS) of Kp (22), including typing methods such as core genome multi-locus sequence typing (cgMLST) (23, 24), has been increasingly used. However, data on Kp transmission and infection apart from outbreak scenarios are still scarce.

Therefore, the objective of our study was to identify patient-related risk factors for nosocomial Kp acquisition in a non-outbreak, tertiary NICU/intermediate care (IMC) setting. The integration of epidemiological Kp surveillance and prospective IPC measures with a routine phenotypic and genomic analysis of Kp isolates, irrespective of resistance profile or outbreak suspicion, enabled us to assess Kp cluster prevalence. Furthermore, we contextualized the genomic features of Kp isolates with public genomes and explored the associations of Kp cluster prevalence with intensive care level and climate conditions using interpretable machine learning.

This comprehensive approach aimed to improve our understanding of the pathogen-patient-environment interactions as a basis for strengthening IPC strategies in the future.

## Methods

### Study type

We conducted a retrospective cohort study of our prospective surveillance program including all patients admitted to the tertiary 21-bed NICU/IMC of Hannover Medical School (MHH) from October 1, 2021, to March 31, 2024. The primary outcome was nosocomial Kp acquisition, defined as detection of Kp on or after day 3 of NICU/IMC stay. The secondary outcome was the prevalence of Kp clusters defined by genomic sequencing. Exploratory analyses compared nosocomial vs community-acquired cases and cluster-assigned vs non-cluster-assigned nosocomial cases. In addition, we characterized phenotypic and genomic features of all Kp isolates. The study adhered to the Strengthening the Reporting of Observational Studies in Epidemiology (STROBE) reporting guideline (25). The ward setting, definitions, and data acquisition are detailed in the Additional Methods (see Additional file 1, Additional Methods). In brief, a patient hospital stay (PHS) was defined as continuous inpatient episode on the NICU/IMC. The time at risk was defined as the number of days from admission to first detection of Kp in any microbiological sample.

### IPC concept and surveillance

Detailed information on our IPC concept, Kp surveillance program, and microbiological and molecular diagnostics are available in the Additional Methods (see Additional file 1, Additional Methods). In short, following microbiological admission screening, all NICU patients and IMC patients with an actual weight <1500g received weekly colonization screening of respiratory secretions and rectal swabs according to KRINKO (20, 21). Microbiological diagnostics were conducted at the ISO 15189-accredited microbiology laboratory of MHH, and each newly emerged Kp isolate was sequenced using Illumina short-read technology.

### Statistical analysis and machine learning

To reduce immortal-time bias, the *at-risk* population for nosocomial Kp acquisition was defined as PHSs lasting ≥48 hours with negative Kp status until that time, resulting in a reduced sample for multivariable regression analyses compared to the full cohort. We performed multivariable logistic regression to estimate adjusted odds ratios (aOR) with 95% profile-likelihood confidence intervals (CIs) for nosocomial acquisition, including gestational age (GA), very low birth weight (<1500g, VLBW, categorical), and sex as covariates (reference categories: absence of VLBW, female sex). Screening effort (number of surveillance samples per patient divided by the individual time at risk) was also included as a covariate to account for potential detection bias. A multivariable Cox proportional hazards model was additionally applied to account for time-dependent effects, with time at risk defined as days from admission to Kp acquisition or discharge (censoring). Covariates were identical to those used in the logistic regression model. Adjusted Hazard ratios (aHRs) with 95% profile-likelihood confidence intervals were estimated. An α level of .05 was used. Epidemiological and clinical variables were summarized as median (interquartile range [IQR]) or number (percentage). Group differences were tested using the chi-square test for categorical and Wilcoxon rank-sum test for continuous variables, with significance at *P*<.05. GraphPad Prism (version 10.2.0; GraphPad Software, Massachusetts, USA) was used for statistical analyses. Reporting of machine learning models followed the TRIPOD+AI statement (26). Detailed information on machine learning analyses is available in the Additional Methods (see Additional file 1, Additional Methods). Machine learning analyses modeled cluster prevalence as the outcome in relation to patient-care metrics and climate factors including 127 weekly observations, with 27 weeks contributing to the test set.

Hyperparameters were tuned to optimize model fit and interpretability, and models were used in a hypothesis-generating rather than predictive capacity. Model hyperparameters are detailed in Additional Table 1.

**Table 1.** Epidemiological and clinical characteristics of all PHSs stratified by Kp status and molecular cluster assignment.

| Parameter | All PHSs | PHSs without Kp (A) | PHSs with any Kp (B) | PHSs with nosocomial Kp (C) | PHSs with nosocomial, cluster-assigned Kp (D) |
| --- | --- | --- | --- | --- | --- |
| **Total PHSs, n (<sup>a</sup>)** | 1009 (936) | 927 (859<sup>b</sup>) | 82 (81) | 58 (57) | 37 (37) |
| **Length of PHSs, median [IQR], days** | 6.0<br>[2.4 – 15.2] | 5.3<br>[2.2 – 13.6] | 28.6<br>[9.1 – 50.7] | 32.8<br>[17.1 – 62.4] | 35.0<br>[26.2 – 57.4] |
| **Time at risk<sup>c</sup>, median [IQR], days** | 6.0<br>[2.4 – 15.2] | 5.3<br>[2.2 – 13.6] | 7.0<br>[2.0 – 18.0] | 12.5<br>[6.8 – 25.5] | 16.0<br>[9.0 – 30.0] |
| **Sex, Male, n (%)** | 523/936 (55.9) | 479/859 (55.8) | 44/81 (54.3) | 32/57 (56.1) | 23/37 (62.2) |
| **VLBW, n (%)** | 202/936 (21.6) | 158/859 (18.4) | 45/81 (55.6) | 38/57 (66.7) | 30/37 (81.1) |
| **GA, median [IQR], days (weeks)** | 254 (36.3)<br>[226 (32.3) – 273 (39.0)] | 256 (36.6)<br>[230 (32.9) – 274 (39.1)] | 219 (31.3)<br>[190 (27.1) – 249 (35.6)] | 208 (29.7)<br>[188 (26.9) – 237 (33.9)] | 202 (28.9)<br>[187 (26.7) – 221 (31.6)] |
| **Birth weight, median [IQR], grams** | 2503<br>[1610 – 3286] | 2600<br>[1718 – 3340] | 1470<br>[960 – 2225] | 1300<br>[955 – 1878] | 1220<br>[930 – 1478] |

Abbreviations and definitions: PHS, patient hospital stay; defined as a continuous inpatient episode on the NICU/IMC. Discharge or transfer ended the PHS, readmission started a new PHS. Kp, *Klebsiella pneumoniae* complex; A Kp case was any PHS with $\geq 1$ Kp-positive specimen. Nosocomial Kp carriage was defined as acquisition on or after day three or later. VLBW, very low birth weight; GA, gestational age. <sup>a</sup> Numbers in parentheses indicate corresponding patients. <sup>b</sup> Four patients acquired Kp during a later PHS. <sup>c</sup> Time at risk is defined as length of stay in the NICU/IMC until Kp acquisition for positive cases or until discharge for negative cases.

## Results

### Epidemiology and primary outcome

During the study period, 936 individual patients (median: 51 patients/month) were admitted to the NICU/IMC, generating 1009 PHSs and 14 684 patient days (median PHS length, 6 days) (**Table 1**). Microbiological diagnostics identified 81 Kp patients (prevalence, 8.7/100 patients; incidence density, 5.6 PHS/1000 patient days). Numbers differ slightly between patients, PHSs, and isolates due to multiple acquisitions in single patients (details in Table footnotes).

The mean (IQR) birth weight of all 936 NICU/IMC patients was 2503 (1610–3286) g, the mean (IQR) GA was 36.3 (32.3 –39.0) weeks. VLBW infants accounted for 21.6% of all patients (202/936) and were threefold more frequent in the Kp subgroup. Among Kp patients, 70.4% (57/81) had nosocomial acquisition, of whom 66.7% (38/57) were VLBW infants. In multivariable logistic regression, VLBW was the only strong risk factor for nosocomial Kp acquisition (aOR, 3.42; 95% CI, 1.29–9.32), whereas higher gestational age was weakly protective (aOR, .98; 95% CI, .97-.9986). After adjustment for screening effort, which was itself strongly associated with Kp acquisition, the association of VLBW remained robust (**Table 2**). In time-to-event analysis accounting for varying time at risk, VLBW infants showed a significantly increased hazard of nosocomial acquisition (aHR 3.49, 95% CI, 1.25-10.08). Exploratory analyses showed that nosocomial cases, compared with community-acquired cases, had a significantly higher proportion of VLBW infants, lower GA, and lower birth weight (Additional Table 2). During the time at risk, nosocomial cases also showed a high proportion of peripheral venous catheters (96.6%), noninvasive ventilation (75.9%), and intravenous antibiotic therapy (43.1%; Additional Table 2).

**Table 2:** Multivariable regression analyses of nosocomial Kp or nosocomial Kp with molecular cluster assignment.

| Parameter | Adjusted OR [95% CI]<sup>a</sup>,<br>nosocomial Kp | Adjusted HR [95% CI]<sup>b</sup>,<br>nosocomial Kp | Adjusted OR [95% CI]<sup>a</sup>,<br>nosocomial and<br>cluster-assigned Kp | Adjusted HR [95% CI]<sup>b</sup>,<br>nosocomial and<br>cluster-assigned Kp |
| --- | --- | --- | --- | --- |
| **Sex** | .84 [.47 – 1.50] | .68 [.39 – 1.16] | .60 [.29 – 1.23] | .51 [.25 – 1.02] |
| **VLBW** | **3.42 [1.29 – 9.32]** | **3.49 [1.25 – 10.08]** | **8.76 [2.45 – 34.16]** | **10.78 [2.61 – 49.69]** |
| **GA** | **.98 [.97 – .9986]** | 1.01 [.99 – 1.02] | .99 [.97 – 1.01] | 1.01 [.99 – 1.03] |
| **Screening effort<sup>c</sup>** | **4.83 [2.01 – 11.10]** | **15.26 [7.64 – 29.11]** | **3.62 [1.05 – 10.78]** | **21.03 [7.63 – 53.59]** |

Abbreviations and definitions: Kp, *Klebsiella pneumoniae* complex; The *at-risk population* analyzed in the regression models comprised patients hospitalized for ≥48 h and Kp negative until that time point (landmark definition; n=659 Kp negative, n=57 nosocomial Kp, and n=37 nosocomial cluster-assigned patients). VLBW, very low birth weight; GA, gestational age. <sup>a</sup>Adjusted odds ratios with 95% profile-likelihood confidence intervals are from multivariable logistic regression at the patient level, comparing nosocomial Kp acquisition or nosocomial Kp with molecular cluster assignment against no Kp detection. Because of collinearity, VLBW (<1500 g birth weight) and not birth weight was retained in the multivariable model. <sup>b</sup>Adjusted hazard ratios with 95% profile-likelihood confidence intervals are from Cox proportional hazards regression based on the same dataset, with individual time at risk, i.e., length of stay in the NICU/IMC until Kp acquisition for positive cases or until discharge for negative cases, as time variable. Kp acquisition was defined as event and discharge without Kp as censor. Reference categories for both analyses: male sex and absence of VLBW. <sup>c</sup>Screening effort was defined as the number of surveillance samples per patient divided by the individual time at risk. Inclusion of screening effort did not materially change the association of VLBW with Kp acquisition in the logistic regression model.

Three patients (3.7%) developed a Kp infection (**Table 3**). One patient developed infection postoperatively following colonization acquired at a transferring hospital, whereas in the two other patients the infection occurred during prolonged hospitalizations; both were profoundly immunocompromised, including one solid organ transplant recipient.

**Table 3:** Epidemiological and clinical characteristics of Kp infections.

| Epidemiological Parameters | Patient A | Patient B | Patient C |
| --- | --- | --- | --- |
| GA, category | Early preterm | Early preterm | Term |
| Birth weight, grams | [1,500-2,499] | [1,500-2,499] | [2,500-4,200] |
| Time at risk<sup>a</sup>, days | 0<sup>b</sup> | 48 | 56 |
| Time until infection, days | 5 | 48 | 56 |
| Length of stay, days | 11 | 86 | 85 |
| **Clinical parameters during time until infection** |  |  |  |
| Central venous catheter, days | 5 | 33 | 56 |
| Peripheral venous catheter, days | 5 | 22 | 5 |
| Invasive ventilation, days | 1 | 14 | 56 |
| Non-invasive ventilation, days | 1 | 11 | 0 |
| Transurethral catheter, days | 0 | 9 | 10 |
| Surgery | yes | no | yes |

Abbreviations and definitions: GA, gestational age; Kp, *Klebsiella pneumoniae* complex; Kp infection was defined as the presence of Kp in a diagnostic microbiological specimen (e.g. blood culture), combined with $\geq 5$ days of antibiotic treatment and clinical documentation.
<sup>a</sup> Time at risk refers to days from admission until acquisition of the Kp isolate. <sup>b</sup> Postoperative infection following Kp colonization at a transferring hospital.

### Nosocomially acquired Kp and molecular cluster assignment

Eighty-three isolates from 79 patients were available for molecular analysis. 40 patients (40/81 [49.4%]) were assigned to 10 genomic clusters, each comprising two to ten patients. Cluster assignment was more frequent among nosocomial Kp patients (37/57 [64.9%]) than among community-acquired Kp patients (3/24 [12.5%]). In multivariable analysis, VLBW was associated with substantially higher odds of cluster assignment (aOR, 8.76; 95% CI, 2.45-34.16; **Table 2**).

Exploratory subgroup analyses showed that nosocomial cluster-assigned cases had more often a VLBW and had higher rates of noninvasive ventilation compared with non-cluster assigned cases (**Table 4**). Moreover, the duration of noninvasive ventilation and peripheral venous catheterization was significantly longer in cluster-assigned cases (Additional Table 3).

**Table 4.**
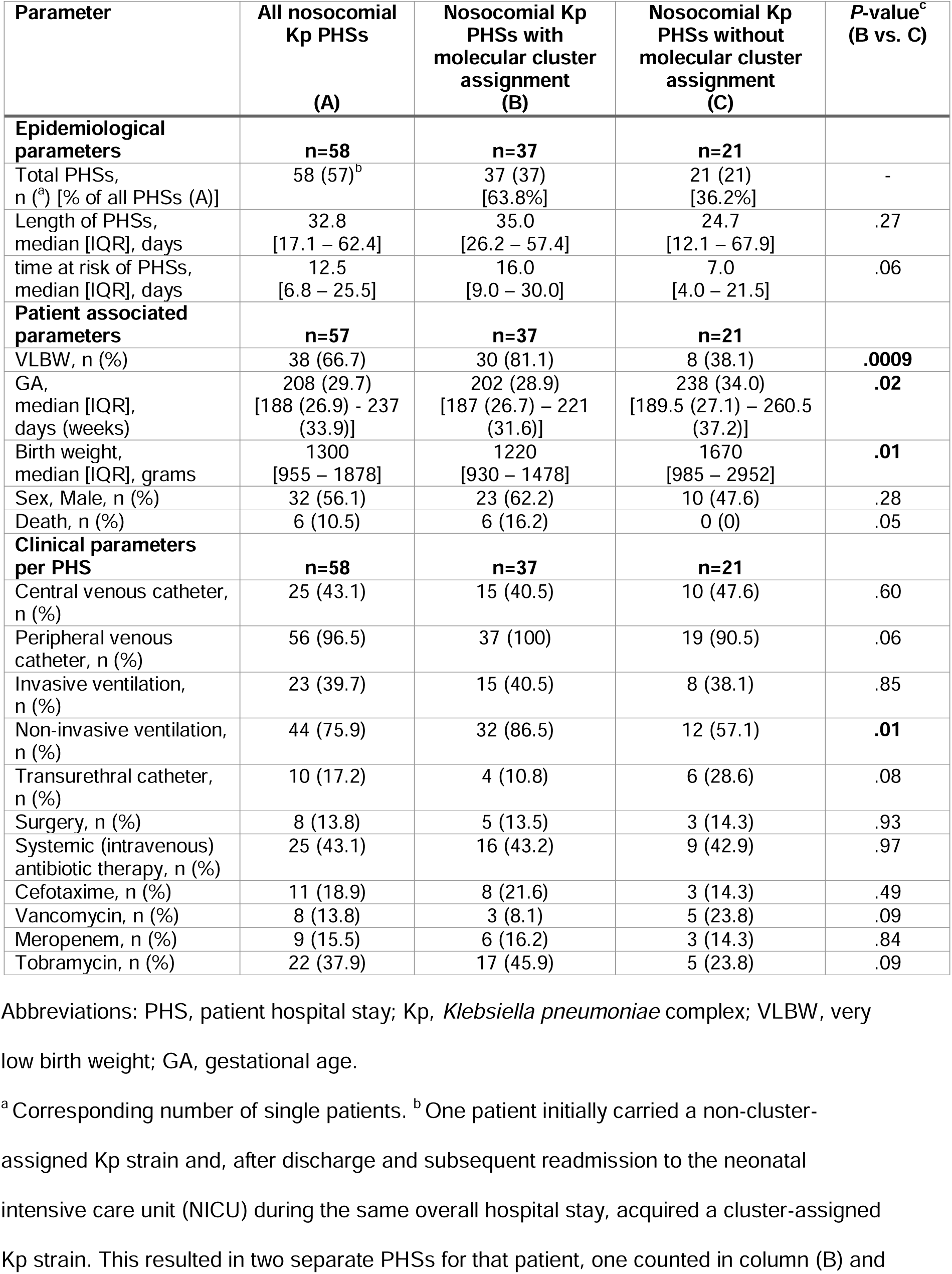
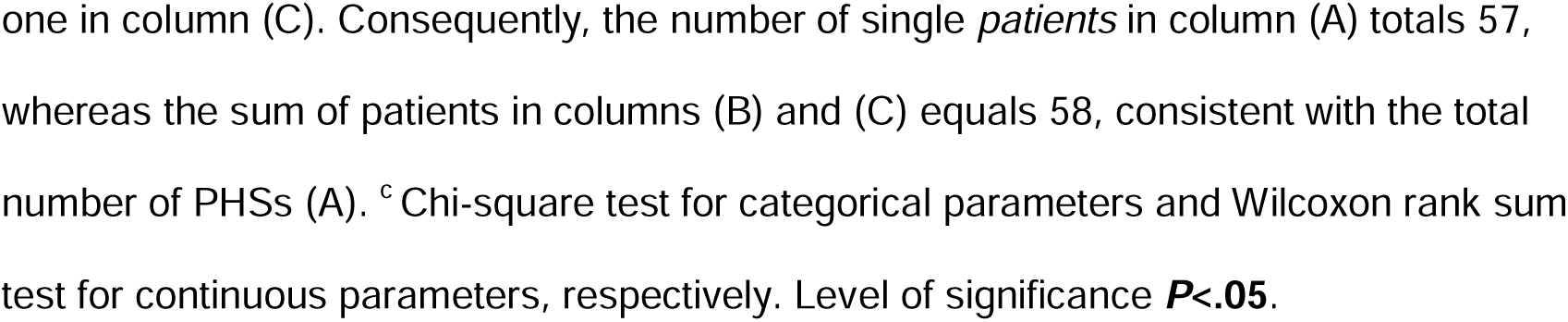
Epidemiological and clinical parameters of nosocomial Kp PHSs stratified by molecular cluster assignment.

| Parameter | All nosocomial Kp PHSs<br>(A) | Nosocomial Kp PHSs with molecular cluster assignment<br>(B) | Nosocomial Kp PHSs without molecular cluster assignment<br>(C) | P-value<sup>c</sup><br>(B vs. C) |
| --- | --- | --- | --- | --- |
| **Epidemiological parameters** | **n=58** | **n=37** | **n=21** |  |
| Total PHSs, n (<sup>a</sup>) [% of all PHSs (A)] | 58 (57)<sup>b</sup> | 37 (37)<br>[63.8%] | 21 (21)<br>[36.2%] | - |
| Length of PHSs, median [IQR], days | 32.8<br>[17.1 – 62.4] | 35.0<br>[26.2 – 57.4] | 24.7<br>[12.1 – 67.9] | .27 |
| time at risk of PHSs, median [IQR], days | 12.5<br>[6.8 – 25.5] | 16.0<br>[9.0 – 30.0] | 7.0<br>[4.0 – 21.5] | .06 |
| **Patient associated parameters** | **n=57** | **n=37** | **n=21** |  |
| VLBW, n (%) | 38 (66.7) | 30 (81.1) | 8 (38.1) | **.0009** |
| GA, median [IQR], days (weeks) | 208 (29.7)<br>[188 (26.9) - 237 (33.9)] | 202 (28.9)<br>[187 (26.7) – 221 (31.6)] | 238 (34.0)<br>[189.5 (27.1) – 260.5 (37.2)] | **.02** |
| Birth weight, median [IQR], grams | 1300<br>[955 – 1878] | 1220<br>[930 – 1478] | 1670<br>[985 – 2952] | **.01** |
| Sex, Male, n (%) | 32 (56.1) | 23 (62.2) | 10 (47.6) | .28 |
| Death, n (%) | 6 (10.5) | 6 (16.2) | 0 (0) | .05 |
| **Clinical parameters per PHS** | **n=58** | **n=37** | **n=21** |  |
| Central venous catheter, n (%) | 25 (43.1) | 15 (40.5) | 10 (47.6) | .60 |
| Peripheral venous catheter, n (%) | 56 (96.5) | 37 (100) | 19 (90.5) | .06 |
| Invasive ventilation, n (%) | 23 (39.7) | 15 (40.5) | 8 (38.1) | .85 |
| Non-invasive ventilation, n (%) | 44 (75.9) | 32 (86.5) | 12 (57.1) | **.01** |
| Transurethral catheter, n (%) | 10 (17.2) | 4 (10.8) | 6 (28.6) | .08 |
| Surgery, n (%) | 8 (13.8) | 5 (13.5) | 3 (14.3) | .93 |
| Systemic (intravenous) antibiotic therapy, n (%) | 25 (43.1) | 16 (43.2) | 9 (42.9) | .97 |
| Cefotaxime, n (%) | 11 (18.9) | 8 (21.6) | 3 (14.3) | .49 |
| Vancomycin, n (%) | 8 (13.8) | 3 (8.1) | 5 (23.8) | .09 |
| Meropenem, n (%) | 9 (15.5) | 6 (16.2) | 3 (14.3) | .84 |
| Tobramycin, n (%) | 22 (37.9) | 17 (45.9) | 5 (23.8) | .09 |

Abbreviations: PHS, patient hospital stay; Kp, *Klebsiella pneumoniae* complex; VLBW, very low birth weight; GA, gestational age.
<sup>a</sup> Corresponding number of single patients. <sup>b</sup> One patient initially carried a non-cluster-assigned Kp strain and, after discharge and subsequent readmission to the neonatal intensive care unit (NICU) during the same overall hospital stay, acquired a cluster-assigned Kp strain. This resulted in two separate PHSs for that patient, one counted in column (B) and
one in column (C). Consequently, the number of single *patients* in column (A) totals 57, whereas the sum of patients in columns (B) and (C) equals 58, consistent with the total number of PHSs (A). ° Chi-square test for categorical parameters and Wilcoxon rank sum test for continuous parameters, respectively. Level of significance ***P* < .05**.

### Integrated phenotypic and genomic analysis

Phenotypic hypervirulence markers yielded one positive string test and two indeterminate tellurite results; none belonged to a genomic cluster or caused infection. Kleborate analysis did not identify hypervirulent strains. Three isolates within cgMLST cluster 8 carried yersiniabactin and colibactin, but none caused infection (Additional File 2, Additional Table 4). Most isolates (63/83) exhibited a wild-type antimicrobial resistance profile; 10 had an extended-spectrum β-lactamase (ESBL) phenotype, of which eight belonged to clusters (ST29 and ST353, four isolates each, all blaCTX-M-15-positive; **Figure 1**). No carbapenem resistance was detected. All cgMLST clusters displayed consistent phenotypic profiles, with a single discrepant isolate in cluster 1 (Split k-mer analysis [SKA] distance >20 single nucleotide polymorphisms [SNPs]). SNP comparison against public genomes (Bacterial and Viral Bioinformatics Resource Center [BV-BRC] database) (27) revealed the closest match (48 SNPs) to a marine isolate from Norway (28); this strain (ST866) was identified upon admission in a patient residing at the German North Sea coast (Additional File 2, Additional Table 5).

**Figure 1.**
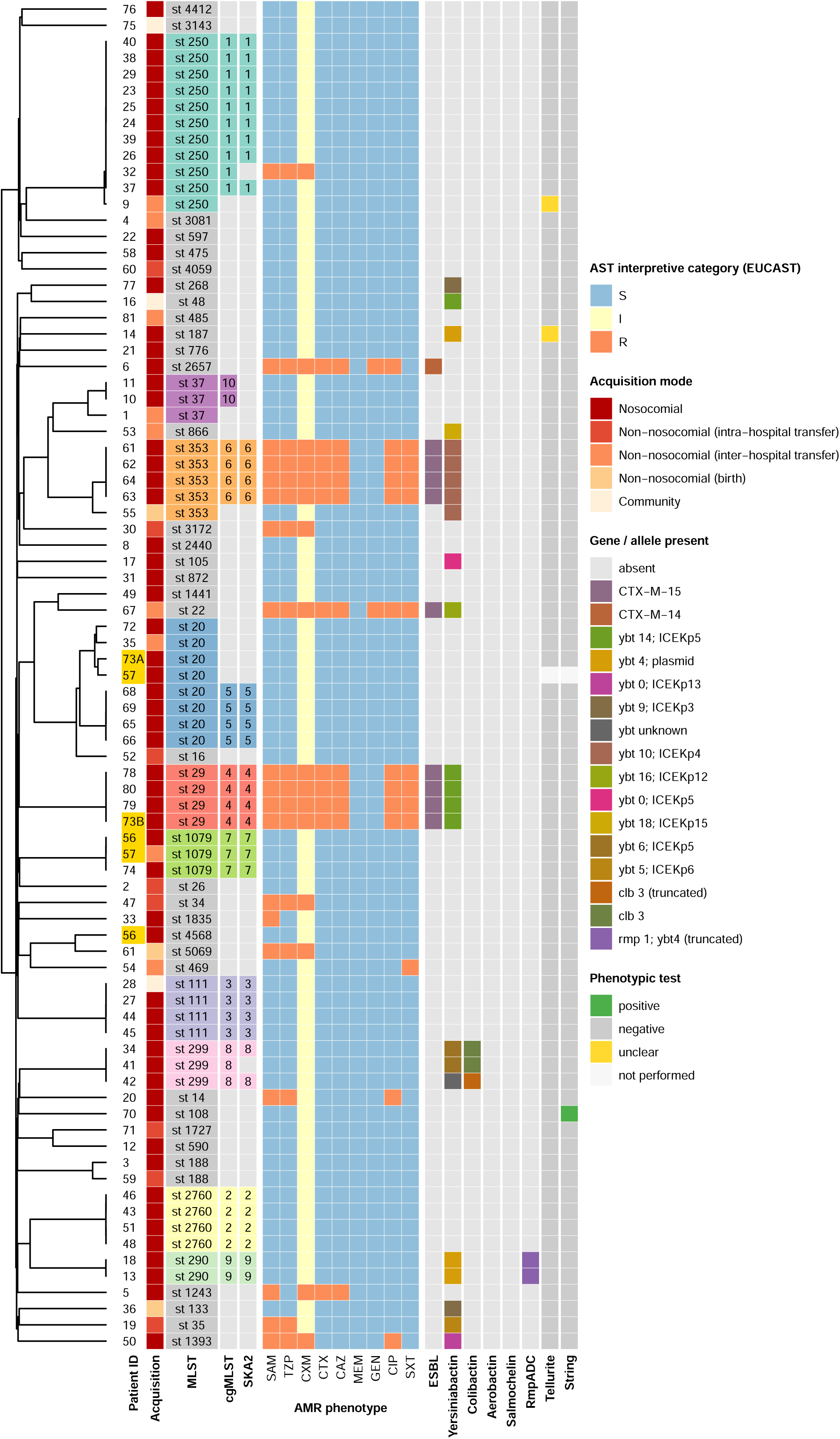
Phylogeny, antimicrobial susceptibility, and genomic/phenotypic features of 83 Kp isolates. Rows represent individual isolates ordered by a midpoint-rooted neighbor-joining dendrogram based on core-genome multilocus sequence typing (cgMLST) allele distances calculated in Ridom SeqSphere+ (left). The central heatmap displays EUCAST antimicrobial susceptibility interpretations (susceptible [S], increased exposure [I], resistant [R]) for nine agents: ampicillin/sulbactam (SAM), piperacillin/tazobactam (TZP), cefuroxime (CXM), cefotaxime (CTX), ceftazidime (CAZ), meropenem (MEM), gentamicin (GEN), ciprofloxacin (CIP), and trimethoprim/sulfamethoxazole (SXT). Left annotations: patient ID (duplicates highlighted), acquisition mode (darkest = nosocomial; lightest = community), and cluster assignment by MLST, cgMLST, and SKA2 (shared color key); unclustered (“singleton”) entries are shown in light gray. Right annotations: presence of extended-spectrum β-lactamase (ESBL) loci and major virulence determinants (Yersiniabactin, Colibactin, Aerobactin, Salmochelin, RmpADC; colored by allele). Phenotypic results for tellurite resistance and string test shown as positive, negative, or unclear; light gray indicates tests not performed.

Throughout the study, molecular typing results were presented during regular interdisciplinary IPC audits. The median time from sampling to completed WGS was 3.1 weeks (IQR, 2.6-4.6).

### Association of climate and patient-care factors with molecular cluster prevalence

Kp cluster prevalence showed seasonal variations (Additional Figure 1). To examine external drivers, machine learning models were trained with climate variables and nurse-to-patient ratios, including 4-week lagged values based on exploratory analyses suggesting temporal associations. Model performance was evaluated using a season-stratified 80:20 split. The reported R² reflects out-of-sample performance on the held-out test set and quantifies the proportion of variance explained within this specific temporal dataset.

Linear models (elastic net, lasso, and ridge) showed limited predictive ability (Additional Table 1). An extreme gradient boosting (XGBoost) model using a reduced feature set provided the best fit to the observed variance (hold-out R² = 0.80). A random forest model achieved a comparable fit (hold-out R² = 0.78) but required inclusion of 24 features (**Figure 2A**; full feature set in Additional file 1, Additional Methods). The largest discrepancy between observed and predicted prevalence occurred in August 2023, mainly attributable to cluster 5 (ST20).

**Figure 2.**
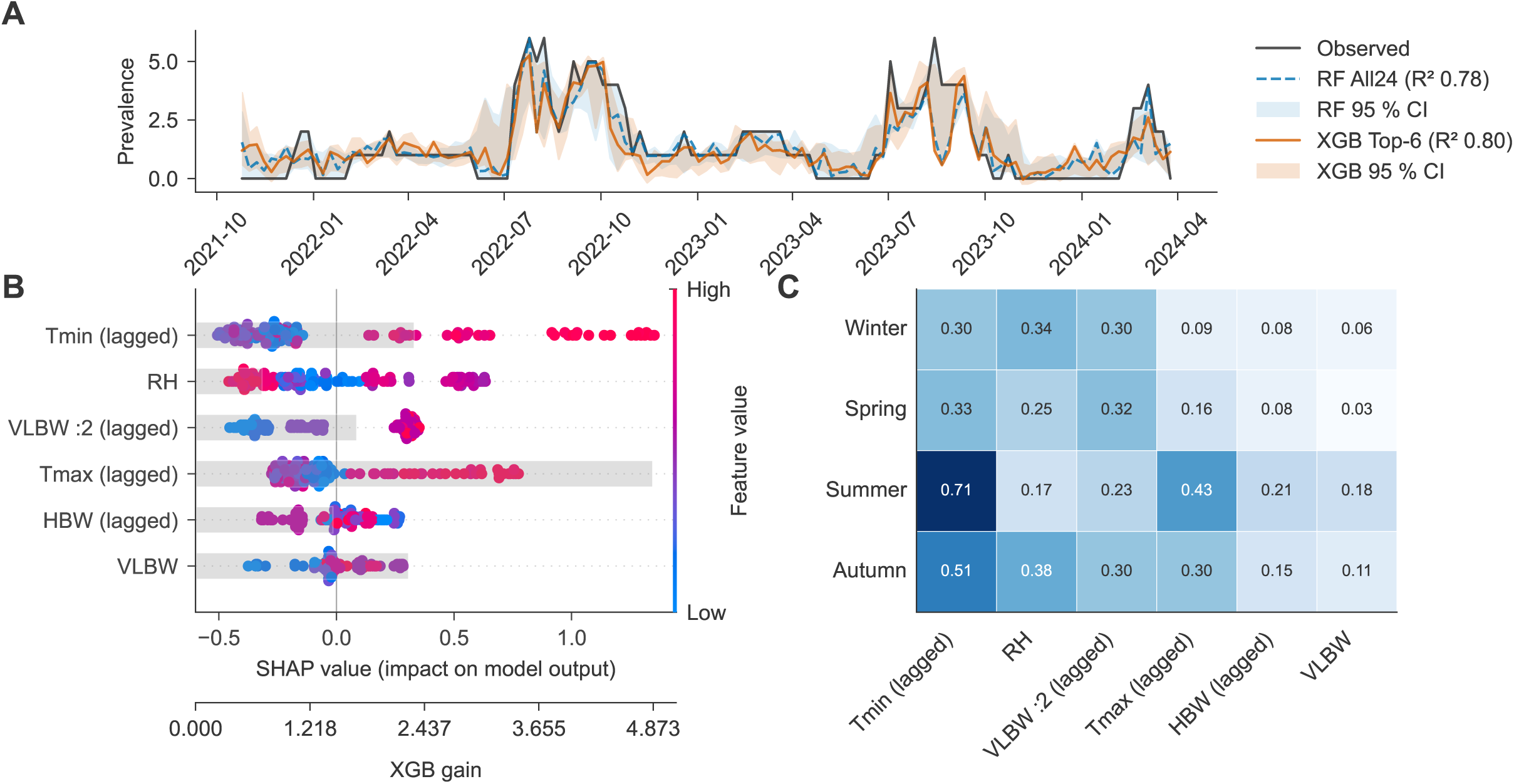
Machine learning-based prediction of molecular cluster prevalence and model interpretation. **(A)** Weekly observed cluster prevalence (black line) vs predicted prevalence from Random Forest (blue dotted line) and reduced-feature extreme gradient boosting (XGBoost) model (orange line). Shaded bands indicate pointwise 95% bootstrap intervals obtained by refitting each model on 800 bootstrap resamples of the weekly dataset and predicting across all weeks. R² values refer to performance on a single within-season 80:20 train-test split. **(B)** Global feature importance in the reduced-feature XGBoost model. Shapley Additive Explanations (SHAP) beeswarm plot shows mean absolute SHAP values (ranked by importance) and individual SHAP values per observation (colored by feature value). Grey bars indicate gain-based feature importance. **(C)** Seasonal variation in feature importance. Heatmap of mean absolute SHAP values stratified by meteorological season. Abbreviations: [Tmin (lagged)] minimum temperature, 4-week lag; [RH] current humidity; [VLBW :2 (lagged)] very low birth weight (<1500g) patients treated with a 1:2 nurse-to-patient ratio (1 nurse per 2 patients), 4-week lag; [Tmax (lagged)] maximum temperature, 4-week lag; [HBW (lagged)] peak number of patients with birth weight >1500g (“higher birth weight”) treated, 4-week lag; [VLBW] current peak number of very low birth weight patients treated (all nursing ratios). For definitions, see Additional Methods.

Feature importance was evaluated using gain-based metrics and Shapley Additive Explanations (SHAP). A higher minimum outdoor temperature four weeks earlier (lagged) consistently emerged as the top feature across seasons – weeks with little or no cooling phases (warmer nights) were followed by higher cluster prevalence. Humidity showed a complex, bidirectional influence on predicted outcomes across its observed range and exerted stronger relative effects in winter and autumn, while the VLBW nurse-to-patient ratio, correlated with higher predicted prevalence, contributed more modestly but stably over time (**Figure 2B-C**, Additional Figure 2). Learning curves demonstrated that cross-validated R² values remained below 0.5 across increasing training fractions, indicating moderate generalizability (Additional Figure 3).

## Discussion

In this study, we comprehensively analyzed epidemiological and genomic Kp surveillance at a tertiary NICU/IMC in a non-outbreak setting, including an assessment of risk factors for nosocomial Kp acquisition and an evaluation of cluster prevalence using machine learning. Kp was detected in ∼8-9% of hospitalized patients, consistent with 2016-2018 data from our unit (29), and about ten times higher than local MRSA prevalence (30). In contrast, intestinal Kp carriage was 40-60% in Indian VLBW preterm infants (31), while adult intensive care unit (ICU) colonization rates ranged between 6-20% (32, 33). More than two thirds of Kp cases in our NICU were nosocomial, consistent with studies showing that the initial microbiome in preterm infants is shaped by the unit’s environment, including other infants, surfaces, staff and parents (34–36). Although VLBW patients accounted for only ∼21% of all PHSs, VLBW was the only independent factor for nosocomial acquisition, with 67% of nosocomial cases and 81% of cluster-associated PHSs occurring in VLBW infants. Time-to-event analyses supported that the association was not solely explained by longer time at risk. Nevertheless, residual confounding by care intensity cannot be fully excluded, and we cannot conclusively distinguish between increased biological susceptibility and increased exposure associated with intensified medical management in VLBW infants.

Kp is a leading cause of neonatal sepsis following intestinal colonization (37–39). Known infection risk factors include patient-related factors, invasive procedures, and prior antibiotic use (40), although pathomechanisms remain elusive (32). We observed increased rates and durations of noninvasive ventilation and peripheral venous catheter use in our nosocomial cluster-associated patients. Peripheral catheters are a recognized risk factor for nosocomial bloodstream infections in VLBW patients (41) and have been addressed in German IPC recommendations (20). However, given the longer time at risk in nosocomial patients, these findings should be interpreted cautiously, as they may reflect opportunity bias.

Most NICU Kp reports focus on multidrug-resistant or hypervirulent strains during outbreaks (12–15, 17). In contrast, our colonization screening explicitly addresses also the carriage of susceptible (wild-type) Kp isolates. No hypervirulent or carbapenem-resistant isolates were identified, suggesting that classical Kp remains the predominant colonizer in NICUs (15, 42, 43). Although phenotypic markers such as string test and tellurite resistance may yield false positives, their use enabled real-time triaging and IPC adjustments before sequencing results were available.

ESBL production was rare and largely confined to two clusters (ST29 and ST353), both harboring blaCTX-M-15, consistent with sporadic import rather than endemic persistence. High genomic consistency within clusters – observed with both cgMLST and SKA in this study supports cgMLST-based IPC interventions. However, the identification of one isolate with divergent resistance within a cgMLST-defined cluster (SNP distance >20) underscores the value of SNP-level analysis to refine cluster boundaries (44). In addition, the literature indicates that, particularly for taxonomically complex species such as Enterobacter cloacae, reliance on cgMLST-based clustering alone may lead to misinterpretation (45). In summary, for the operational use of genomic data in the IPC context, we consider it advisable to interpret cgMLST-based cluster assignments in close conjunction with the epidemiological context and, particularly in cases of uncertainty, to complement them with higher-resolution analytical approaches such as SKA. Interestingly, the closest genomic match for one isolate (ST866) was a marine environmental strain from Norway, illustrating potential environmental influx and the value of contextualizing local isolates against public datasets.

While the spread of hospital-associated, carbapenem-resistant lineages of Kp is well-documented (46), we found no clusters involving globally circulating lineages. Consistent with the absence of hvKp/carbapenem resistance and rapid containment, the infection attack rate of 3.7% of all Kp patients (0.3% of admissions) was comparable to prior reports (6, 39).

Kp was repeatedly introduced and also transmitted, but cluster size remained small (median, four patients), suggesting that transmission was usually limited in size and duration, consistent with timely IPC interventions. These included training/feedback, cohorting/isolation, encouraging hand hygiene, intensified patient and environmental screening, and enhanced disinfection (12–15, 17). Audit-style feedback on epidemiological and genomic findings was progressively strengthened during the study, reaching near-weekly intervals, with rising interdisciplinary participation. Similar formats have proven effective in teaching IPC (47). Clusters detected by molecular typing were addressed systematically during audits and in scheduled or ad hoc interdisciplinary training sessions, prompting targeted interventions to interrupt transmission chains and prevent establishment of dominant ‘ward clones’.

Six pairs of twins were included among cluster patients, highlighting elevated risks for patient-to-patient or parent-to-patient transmission (35, 36). Though skin-to-skin (kangaroo) care in VLBW infants facilitates bacterial exchange, it remains a standard practice that favors neurodevelopment and microbiome stability (48, 49). From an IPC perspective, awareness of this potential transmission route in preterm multiples is essential.

Seasonal variation in Kp cluster prevalence, despite stable NICU occupancy, prompted further investigation using machine learning models. Tree-based models, particularly a reduced-feature XGBoost model, explained up to 80% of variance in weekly prevalence within the observed dataset, while linear models failed to capture nonlinear relationships. These findings suggest that Kp transmission in NICUs is shaped not only by patient-specific factors, but also by structural and environmental variables with delayed effects.

A higher minimum daily outdoor temperature four weeks earlier was the most influential predictor, consistent with multicenter studies linking temperature with bloodstream infection incidence and environmental Kp abundance (50–53). In particular, Schwab *et al.* reported that nosocomial bloodstream infection rates in German ICUs were approximately 16% higher in months with mean temperatures ≥20 °C compared with <5 °C, with the strongest association observed for temperatures in the preceding month (50). Additional drivers in our data included relative humidity and increased intensive-care density/capacity, with the VLBW nurse-to-patient ratio serving as a surrogate for patient-related risk. We hypothesize that climate conditions may influence staff and visitor colonization on one hand and accumulation of Kp in the ward environment (e.g., surfaces, devices, sinks, wastewater systems, or other moist reservoirs) on the other hand. The importance of lagged features appears biologically and operationally plausible. The lag of four weeks could reflect gradual amplification in environmental burden followed by increased exposure risk and subsequent transmission within the ward setting. The lag may also correspond to the interval between patient acquisition event and eventual microbiological detection. This interval could encompass asymptomatic colonization, intra-ward spread, and eventual sampling.

These models were exploratory and used to quantify associations, not to predict future risk. The identified associations between macroclimatic variables and cluster prevalence should not be interpreted as direct mechanistic effects but rather as statistical relationships within the observed dataset that warrant further investigation in larger multicenter cohorts incorporating environmental measurements. Moreover, the process of variable/feature selection probably influenced model performance and effect estimates, particularly in our study with a limited number of temporal observations. Nonetheless, the models offer a framework for hypothesis generation and provide actionable insights for IPC planning, such as anticipating periods of increased transmission risk. Nonlinear relationships, such as the bidirectional effect of humidity, underline the complexity of these dynamics. Expansion of an ST20 cluster despite predictively unfavorable external influences may suggest clone-specific transmissibility, although we could not find literature specifically supporting this.

This study has several limitations that should be considered when interpreting the findings. This single-center retrospective cohort study includes weekly colonization screening mandated by German IPC regulations, which may increase awareness of bacterial carriage and antibiotic use in preterm infants. Therefore, it may not be directly generalizable to other NICU settings. Routine Kp sequencing was subject to a time lag, limiting its utility for immediate IPC interventions. While we adjusted the primary analysis to reduce immortal time bias, differences in observation periods between nosocomial and community-acquired cases may have introduced opportunity bias. The machine learning models were not externally validated, and prone to overfitting, requiring larger multicenter datasets for predictive use.

The R² estimate reported reflects model fit within 127 weekly observations of a single-center cohort and should be interpreted as descriptive within this specific temporal context rather than as evidence of robust predictive performance across settings. The study period was relatively short for drawing climate-related conclusions. In addition, the effect of daily outdoor temperature, rather than indoor temperature, on cluster prevalence was analyzed. Practical observations on the ward suggest that, despite the presence of ventilation systems, outdoor temperature affected indoor thermal conditions. Nonetheless, the absence of indoor environmental measurements further limits the ability to infer any causal pathways. In sum, the observed associations should therefore be interpreted as hypothesis-generating rather than confirmatory evidence of climate-driven transmission dynamics. Additional potential transmission vectors – such as staff and visitor carriage or environmental reservoirs – were not systematically assessed.

Because molecular results were delayed, findings mainly informed medium- to long-term IPC strategies in our setting. Streamlined workflows (e.g. usage of Oxford Nanopore Sequencing technologies (54, 55) or more rapid Illumina sequencing) may allow more actionable short-term interventions in the future.

## Conclusions

In a tertiary NICU/IMC, prospective genomic surveillance within a structured IPC program showed that Kp transmission was mainly driven by wild-type isolates. Infection rates were uncommon and genomic clusters remained small, consistent with the effect of routine screening and sequencing-informed audits with rapid feedback into practice. VLBW infants were the main risk group for nosocomial acquisition and were disproportionately affected by cluster-associated Kp. Cluster prevalence reflected the combined influence of VLBW patient vulnerability, structural, and environmental influences, with higher rates observed following periods of sustained warmer nights. These findings outline a scalable surveillance-to-action approach that may help further limit transmission in highly vulnerable neonatal populations. The identified time lag suggests a planning horizon for risk-triggered IPC: timely staff refreshers, increased environmental disinfection, focused screening, and the preemptive reverse isolation of infants at highest risk during anticipated higher-risk periods could be measures worth evaluating their efficacy in future studies.

## Data Availability

Patient data used in this study are confidential in accordance with the German Data Privacy Act, the ethics committee, and the data protection commissioner of Hannover Medical School. Patient-related information such as ward of admission, age, sex, underlying disease, or length of stay constitutes indirect identifiers that could potentially allow re-identification. To ensure confidentiality and protect privacy, only anonymized and aggregated data are available for research use, in compliance with the Data Privacy Act. Interested researchers may contact the corresponding authors to request access to anonymized data, subject to approval by the data protection commissioner of Hannover Medical School. All isolate-level metadata used in this study are provided in the Online Supplement (eTable 4-5 in **Supplement 1**). Genomic data have been deposited in Pathogenwatch (https://pathogen.watch); accessions (Genome IDs) are listed in eTable 4 in **Supplement 1**.

## List of abbreviations (alphabetical order)

aHR: adjusted hazard ratio
aOR: adjusted odds ratio
BV-BRC: Bacterial and Viral Bioinformatics Resource Center
cgMLST: core genome multi-locus sequence typing
CI: confidence interval
ESBL: extended-spectrum β-lactamase
GA: gestational age
hvKp: hypervirulent *Klebsiella pneumoniae* complex
ICU: intensive care unit
IMC: intermediate care unit
IPC: infection prevention and control
Kp: *Klebsiella pneumoniae* complex
KRINKO: Commission for Infection Prevention and Hygiene in Healthcare and Nursing at the Robert Koch Institute, Germany (*Kommission für Infektionsprävention in medizinischen Einrichtungen und in Einrichtungen und Unternehmen der Pflege und Eingliederungshilfe*)
MHH: Hannover Medical School (Medizinische Hochschule Hannover)
MRSA: Methicillin-resistant *Staphylococcus aureus*
NICU: neonatal intensive care unit
PHS: patient hospital stay
SHAP: Shapley Additive Explanations
SKA: split k-mer analysis
SNP: single nucleotide polymorphism
VLBW: very low birth weight
VRE: Vancomycin-resistant enterococci
WGS: whole genome sequencing
XGBoost: extreme gradient boosting

## Declarations

### Ethics approval and consent to participate

The institutional review board (ethics committee) of Hannover Medical School approved this study (10682_BO_K_2022). Given that this was a retrospective, quality-assuring study, the need for informed consent was waived by the Ethics Committee and the Data Protection Commissioner of Hannover Medical School.

### Consent for publication

Not applicable.

### Availability of data and materials

The patient data used in this study are confidential in accordance with the German Data Privacy Act, the ethics committee and the data protection commissioner of Hannover Medical School. Patient-related data such as date of ward admission, age, sex, underlying disease or length of stay are indirect identifiers that might enable patient identification. To protect patient confidentiality and participant privacy, the data used for this study can be obtained in anonymized and condensed form only, according to the Data Privacy Act. Interested researchers who meet the criteria for access to confidential data may contact the data protection commissioner of the Hannover Medical School and one of the corresponding authors (e.g.) to obtain access to anonymized data, as approved by the data protection commissioner and the ethics committee of the Hannover Medical School.

All isolate-level metadata used in this study are provided in the Additional File 2 (Additional Tables 4-5). Whole-genome sequencing data have been deposited in the European Nucleotide Archive under study accession **PRJEB98673**; individual sample accession numbers are listed in Additional Table 4.

### Competing interests

The authors declare that they have no competing interests.

### Funding

This research did not receive any specific grant from funding agencies in the public, commercial, or not-for-profit sectors.

### Authors’ contributions

CBO, CB, LK: conceptualization, methodology, data acquisition, formal analysis, validation, writing original draft and review and editing

JE, ML: formal analysis, validation, draft review and editing

CZ: data acquisition, validation, draft review and editing

EE, DS: validation, draft review and editing

SP, CP, BB: validation, draft review and editing

## Acknowledgements

The authors thank all technical staff of the medical microbiology and hospital hygiene laboratories involved in bacterial isolation, phenotypic virulence testing, strain collection, and library preparation. In addition, the authors would like to acknowledge Dr. Ludwig Sedlacek and PD Dr. Marius Vital, who were supervising the routine sequencing workflow in the medical microbiology laboratory. Furthermore, the authors are very grateful to all clinical staff and participating families who contributed to this study.

## Notes

### Competing Interest Statement

The authors have declared no competing interest.

### Funding Statement

This study did not receive any external funding.

### Author Declarations

Ethics committee of Hannover Medical School gave ethical approval for this work (numbers 9365_BO_K_2020 and 10372_BO_K_2022). Ethics committee of Hannover Medical School waived the need for informed consent.

### Summary of Updates

We performed an additional multivariable Cox proportional hazards regression analysis using time at risk as the time-to-event variable and adjusting for gestational age, sex, and screening effort. In this time-dependent model, VLBW remained independently associated with nosocomial acquisition (aHR 3.49, 95% CI, 1.25-10.08, p = 0.019), indicating that the association cannot be explained solely by longer exposure time. We have now clarified in the revised manuscript that the reported R2 of 0.80 reflects the model fit within the observed dataset and should be interpreted as descriptive rather than as evidence of stable predictive performance beyond the study period. Furthermore, we have revised the wording throughout the manuscript to avoid any implication of a direct causal relationship. Specifically, we replaced formulations that could be interpreted as mechanistic or causal with more cautious language emphasizing association and statistical explanation of variance. We phrased some limitations of statistical or ML methods more explicitly and expanded the explanation of our rationale for feature selection in the Additional Methods.

## References

1. Moussa B, Oumokhtar B, Arhoune B, Massik A, Elfakir S, Khalis M, et al. Gut acquisition of Extended-spectrum β-lactamases-producing Klebsiella pneumoniae in preterm neonates: Critical role of enteral feeding, and endotracheal tubes in the neonatal intensive care unit (NICU). PLoS One. 2023;18(11):e0293949.

2. Magill SS, Edwards JR, Bamberg W, Beldavs ZG, Dumyati G, Kainer MA, et al. Multistate point-prevalence survey of health care-associated infections. N Engl J Med. 2014;370(13):1198–208.

3. Vogiantzi G, Metallinou D, Tigka M, Deltsidou A, Nanou CI. Bloodstream Infections in the Neonatal Intensive Care Unit: A Systematic Review of the Literature. Cureus. 2024;16(8):e68057.

4. Sampah MES, Hackam DJ. Dysregulated Mucosal Immunity and Associated Pathogeneses in Preterm Neonates. Front Immunol. 2020;11:899.

5. Humberg A, Fortmann I, Siller B, Kopp MV, Herting E, Göpel W, et al. Preterm birth and sustained inflammation: consequences for the neonate. Semin Immunopathol. 2020;42(4):451–68.

6. Wei X, Liang J, Zhang H, Yan C, Lu X, Chen Y, et al. Clinical features and risk factors of Klebsiella pneumoniae infection in premature infants: a retrospective cohort study. BMC Infect Dis. 2024;24(1):1311.

7. de Lajartre OB, Maamar A, Dejoies L, Delamaire F. Community-acquired hypervirulent Klebsiella pneumoniae invasive infection in critically ill patients who dramatically improved. Eur J Clin Microbiol Infect Dis. 2022;41(10):1275–7.

8. Lam MMC, Wick RR, Watts SC, Cerdeira LT, Wyres KL, Holt KE. A genomic surveillance framework and genotyping tool for Klebsiella pneumoniae and its related species complex. Nat Commun. 2021;12(1):4188.

9. Watanabe N, Masuda A, Watari T, Otsuka Y, Yamagata K, Fujioka M. Evaluation of an optimal agar medium for detecting hypervirulent Klebsiella pneumoniae using string test. Access Microbiol. 2024;6(9).

10. Wu X, Zhan F, Zhang J, Chen S, Yang B. Identification of hypervirulent Klebsiella pneumoniae carrying terW gene by MacConkey-potassium tellurite medium in the general population. Front Public Health. 2022;10:946370.

11. Lan P, Jiang Y, Zhou J, Yu Y. A global perspective on the convergence of hypervirulence and carbapenem resistance in Klebsiella pneumoniae. J Glob Antimicrob Resist. 2021;25:26–34.

12. Shi Q, Zhao J, Wei L, Zhu F, Ji J, Meng Y, et al. Transmission of ST45 and ST2407 extended-spectrum β-lactamase-producing Klebsiella pneumoniae in neonatal intensive care units, associated with contaminated environments. J Glob Antimicrob Resist. 2022;31:309–15.

13. Baek EH, Kim SE, Kim S, Lee S, Cho OH, In Hong S, et al. Successful control of an extended-spectrum beta-lactamase-producing Klebsiella pneumoniae ST307 outbreak in a neonatal intensive care unit. BMC Infect Dis. 2020;20(1):166.

14. Priante E, Minotti C, Contessa C, Boschetto M, Stano P, Dal Bello F, et al. Successful Control of an Outbreak by Phenotypically Identified Extended-Spectrum Beta-Lactamase-Producing Klebsiella pneumoniae in a Neonatal Intensive Care Unit. Antibiotics (Basel). 2022;11(11).

15. Mavroidi A, Liakopoulos A, Gounaris A, Goudesidou M, Gaitana K, Miriagou V, et al. Successful control of a neonatal outbreak caused mainly by ST20 multidrug-resistant SHV-5-producing Klebsiella pneumoniae, Greece. BMC Pediatr. 2014;14:105.

16. Wisgrill L, Lepuschitz S, Blaschitz M, Rittenschober-Böhm J, Diab-El Schahawi M, Schubert S, et al. Outbreak of Yersiniabactin-producing Klebsiella pneumoniae in a Neonatal Intensive Care Unit. Pediatr Infect Dis J. 2019;38(6):638–42.

17. Pathak A, Tejan N, Dubey A, Chauhan R, Fatima N, Jyoti, et al. Outbreak of colistin resistant, carbapenemase (bla (NDM), bla (OXA-232)) producing Klebsiella pneumoniae causing blood stream infection among neonates at a tertiary care hospital in India. Front Cell Infect Microbiol. 2023;13:1051020.

18. Magobo RE, Ismail H, Lowe M, Strasheim W, Mogokotleng R, Perovic O, et al. Outbreak of NDM-1- and OXA-181-Producing Klebsiella pneumoniae Bloodstream Infections in a Neonatal Unit, South Africa. Emerg Infect Dis. 2023;29(8):1531–9.

19. Li J, Shen L, Qian K. Global, regional, and national incidence and mortality of neonatal sepsis and other neonatal infections, 1990-2019. Front Public Health. 2023;11:1139832.

20. KRINKO. Praktische Umsetzung sowie krankenhaushygienische und infektionspra□ventive Konsequenzen des mikrobiellen Kolonisationsscreenings bei intensivmedizinisch behandelten Fru□h- und Neugeborenen Erga□nzende Empfehlung der KRINKO beim Robert Koch-Institut, Berlin, zur Implementierung der Empfehlungen zur Pra□vention nosokomialer Infektionen bei neonatologischen Intensivpflegepatienten mit einem Geburtsgewicht unter 1.500 g aus dem Jahr 2007 und 2012. Epidem Bull. 2013;42:421–33.

21. Litz JE, Goedicke-Fritz S, Härtel C, Wagenpfeil G, Zemlin M, Simon A. Umsetzung des mikrobiologischen Kolonisationsscreenings: Umfrage an 80 neonatologischen Intensivstationen. Epid Bull. 2019;37:387 – 92.

22. Carlos CC, Masim MAL, Lagrada ML, Gayeta JM, Macaranas PKV, Sia SB, et al. Genome Sequencing Identifies Previously Unrecognized Klebsiella pneumoniae Outbreaks in Neonatal Intensive Care Units in the Philippines. Clin Infect Dis. 2021;73(Suppl_4):S316–s24.

23. Gona F, Comandatore F, Battaglia S, Piazza A, Trovato A, Lorenzin G, et al. Comparison of core-genome MLST, coreSNP and PFGE methods for Klebsiella pneumoniae cluster analysis. Microb Genom. 2020;6(4).

24. Hallbäck ET, Johnning A, Myhrman S, Studahl M, Hentz E, Elfvin A, et al. Outbreak of OXA-48-producing Enterobacteriaceae in a neonatal intensive care unit in Western Sweden. Eur J Clin Microbiol Infect Dis. 2023;42(5):597–605.

25. von Elm E, Altman DG, Egger M, Pocock SJ, Gøtzsche PC, Vandenbroucke JP. The Strengthening the Reporting of Observational Studies in Epidemiology (STROBE) statement: guidelines for reporting observational studies. Ann Intern Med. 2007;147(8):573–7.

26. Collins GS, Moons KGM, Dhiman P, Riley RD, Beam AL, Van Calster B, et al. TRIPOD+AI statement: updated guidance for reporting clinical prediction models that use regression or machine learning methods. BMJ. 2024;385:e078378.

27. Olson RD, Assaf R, Brettin T, Conrad N, Cucinell C, Davis JJ, et al. Introducing the Bacterial and Viral Bioinformatics Resource Center (BV-BRC): a resource combining PATRIC, IRD and ViPR. Nucleic Acids Res. 2023;51(D1):D678–d89.

28. Håkonsholm F, Hetland MAK, Svanevik CS, Lunestad BT, Löhr IH, Marathe NP. Insights into the genetic diversity, antibiotic resistance and pathogenic potential of Klebsiella pneumoniae from the Norwegian marine environment using whole-genome analysis. Int J Hyg Environ Health. 2022;242:113967.

29. Baier C, Pirr S, Ziesing S, Ebadi E, Hansen G, Bohnhorst B, et al. Prospective surveillance of bacterial colonization and primary sepsis: findings of a tertiary neonatal intensive and intermediate care unit. J Hosp Infect. 2019;102(3):325–31.

30. Böhne C, Knegendorf L, Schwab F, Ebadi E, Bange FC, Vital M, et al. Epidemiology and infection control of Methicillin-resistant Staphylococcus aureus in a German tertiary neonatal intensive and intermediate care unit: A retrospective study (2013-2020). PLoS One. 2022;17(9):e0275087.

31. Sharma N, Chaudhry R, Panigrahi P. Quantitative and qualitative study of intestinal flora in neonates. J Glob Infect Dis. 2012;4(4):188–92.

32. Martin RM, Cao J, Brisse S, Passet V, Wu W, Zhao L, et al. Molecular Epidemiology of Colonizing and Infecting Isolates of Klebsiella pneumoniae. mSphere. 2016;1(5).

33. Gorrie CL, Mirceta M, Wick RR, Edwards DJ, Thomson NR, Strugnell RA, et al. Gastrointestinal Carriage Is a Major Reservoir of Klebsiella pneumoniae Infection in Intensive Care Patients. Clin Infect Dis. 2017;65(2):208–15.

34. Brooks B, Olm MR, Firek BA, Baker R, Geller-McGrath D, Reimer SR, et al. The developing premature infant gut microbiome is a major factor shaping the microbiome of neonatal intensive care unit rooms. Microbiome. 2018;6(1):112.

35. Brooks B, Firek BA, Miller CS, Sharon I, Thomas BC, Baker R, et al. Microbes in the neonatal intensive care unit resemble those found in the gut of premature infants. Microbiome. 2014;2(1):1.

36. Conceição T, Aires de Sousa M, Miragaia M, Paulino E, Barroso R, Brito MJ, et al. Staphylococcus aureus reservoirs and transmission routes in a Portuguese Neonatal Intensive Care Unit: a 30-month surveillance study. Microb Drug Resist. 2012;18(2):116–24.

37. Schwartz DJ, Shalon N, Wardenburg K, DeVeaux A, Wallace MA, Hall-Moore C, et al. Gut pathogen colonization precedes bloodstream infection in the neonatal intensive care unit. Sci Transl Med. 2023;15(694):eadg5562.

38. Moftian N, Rezaei-Hachesu P, Arab-Zozani M, Samad-Soltani T, Esfandiari A, Tabib MS, et al. Prevalence of gram-negative bacteria and their antibiotic resistance in neonatal sepsis in Iran: a systematic review and meta-analysis. BMC Infect Dis. 2023;23(1):534.

39. Reichert F, Piening B, Geffers C, Gastmeier P, Bührer C, Schwab F. Pathogen-Specific Clustering of Nosocomial Blood Stream Infections in Very Preterm Infants. Pediatrics. 2016;137(4).

40. Olsen AL, Reinholdt J, Jensen AM, Andersen LP, Jensen ET. Nosocomial infection in a Danish Neonatal Intensive Care Unit: a prospective study. Acta Paediatr. 2009;98(8):1294–9.

41. Geffers C, Gastmeier A, Schwab F, Groneberg K, Rüden H, Gastmeier P. Use of central venous catheter and peripheral venous catheter as risk factors for nosocomial bloodstream infection in very-low-birth-weight infants. Infect Control Hosp Epidemiol. 2010;31(4):395–401.

42. Mukherjee S, Mitra S, Dutta S, Basu S. Neonatal Sepsis: The Impact of Carbapenem-Resistant and Hypervirulent Klebsiella pneumoniae. Front Med (Lausanne). 2021;8:634349.

43. Hu Y, Yang Y, Feng Y, Fang Q, Wang C, Zhao F, et al. Prevalence and clonal diversity of carbapenem-resistant Klebsiella pneumoniae causing neonatal infections: A systematic review of 128 articles across 30 countries. PLoS Med. 2023;20(6):e1004233.

44. Wang J, Feng Y, Zong Z. Worldwide transmission of ST11-KL64 carbapenem-resistant Klebsiella pneumoniae: an analysis of publicly available genomes. mSphere. 2023;8(4):e0017323.

45. Danjean M, Royer G, Rodriguez C, Woerther PL, Decousser JW. Genomic pitfalls in hospital surveillance: unexpected Enterobacter cloacae clustering without epidemiological link highlights the need for cautious core genome MLST analysis interpretation. Antimicrob Resist Infect Control. 2025;14(1):152.

46. David S, Reuter S, Harris SR, Glasner C, Feltwell T, Argimon S, et al. Epidemic of carbapenem-resistant Klebsiella pneumoniae in Europe is driven by nosocomial spread. Nat Microbiol. 2019;4(11):1919–29.

47. Mohammedi SB, Gillois P, Landelle C. Nursing students’knowledge and effectiveness of teaching in infection prevention and control. BMC Nurs. 2025;24(1):850.

48. Altit G, Hamilton D, O’Brien K. Skin-to-skin care (SSC) for term and preterm infants. Paediatr Child Health. 2024;29(4):238–54.

49. Hendricks-Muñoz KD, Xu J, Parikh HI, Xu P, Fettweis JM, Kim Y, et al. Skin-to-Skin Care and the Development of the Preterm Infant Oral Microbiome. Am J Perinatol. 2015;32(13):1205–16.

50. Schwab F, Gastmeier P, Hoffmann P, Meyer E. Summer, sun and sepsis-The influence of outside temperature on nosocomial bloodstream infections: A cohort study and review of the literature. PLoS One. 2020;15(6):e0234656.

51. Anderson DJ, Richet H, Chen LF, Spelman DW, Hung YJ, Huang AT, et al. Seasonal variation in Klebsiella pneumoniae bloodstream infection on 4 continents. J Infect Dis. 2008;197(5):752–6.

52. Eber MR, Shardell M, Schweizer ML, Laxminarayan R, Perencevich EN. Seasonal and temperature-associated increases in gram-negative bacterial bloodstream infections among hospitalized patients. PLoS One. 2011;6(9):e25298.

53. Aloufi AS. MaxEnt modeling of Klebsiella pneumoniae: predicting future distribution and evaluating the risk for public health. Geomatics, Natural Hazards and Risk. 2024;15(1).

54. Foster-Nyarko E, Cottingham H, Wick RR, Judd LM, Lam MMC, Wyres KL, et al. Nanopore-only assemblies for genomic surveillance of the global priority drug-resistant pathogen, Klebsiella pneumoniae. Microb Genom. 2023;9(2).

55. Wagner GE, Dabernig-Heinz J, Lipp M, Cabal A, Simantzik J, Kohl M, et al. Real-Time Nanopore Q20+ Sequencing Enables Extremely Fast and Accurate Core Genome MLST Typing and Democratizes Access to High-Resolution Bacterial Pathogen Surveillance. J Clin Microbiol. 2023;61(4):e0163122.

