## Additional File 1 for "Prospective genomic and epidemiologic surveillance of *Klebsiella pneumoniae* in a tertiary NICU"

**Additional Methods.**

**Additional Table 1.** Hold-out performances and hyperparameters of trained machine-learning models.

**Additional Table 2.** Epidemiological and clinical parameters during time at risk of PHSs with nosocomial and community-acquired Kp.

**Additional Table 3.** Duration of clinical parameters in nosocomial Kp PHSs stratified by molecular cluster assignment.

**Additional Table 4.** Isolate-level pheno-genomic metadata, patient/PHS linkage, and Pathogenwatch/Kleborate outputs.

**Additional Table 5.** SKA2 SNP distances between study *Klebsiella pneumoniae* sensu stricto genomes and BV-BRC public genomes, with accessions and isolation metadata.

**Additional Figure 1.** Weekly cluster prevalence and concurrent climate and patient-care load.

**Additional Figure 2.** SHAP dependence plots of key features in the reduced-feature XGBoost model.

**Additional Figure 3.** Learning curves of the reduced-feature XGBoost model demonstrating model performance as a function of training dataset size.

**Additional References.**

### Additional Methods.

### Setting

The combined neonatal intensive care unit (NICU)/intermediate care unit (IMC) provided 21 beds for extremely and very low birth weight (VLBW) preterm infants, and term neonates. The ward comprised 12 intensive care and 9 IMC beds, with intensive care beds being located in five rooms of one to four beds, and the IMC beds being divided into three rooms. Sections of the ward were equipped with a ventilation system, with ICU rooms featuring a three-stage filtration. Nurse-to-patient ratios ranged from 1:1 to 1:4 based on German guidelines for the required standard of care[1]. In detail, standard nurse-to-patient ratio at a tertiary German NICU presumes a 1:2 nurse-to-patient level of care. This level of care applies to cardiorespiratory stable patients who require, i.e., non-invasive ventilation, central venous catheters, thoracic drainages, receive continuous administration of sedatives or catecholamines, or require infection prevention and control (IPC) cohorting measures. A 1:1 nurse-to-patient ratio was implemented for extremely low birth weight infants (<1000g) during their first three days of life, cardiorespiratory unstable, invasively ventilated patients, patients with invasive arterial blood pressure monitoring, patients undergoing hypothermia treatment, isolated patients with single-room assignment, patients at the day of an operation, or during end-of-life care. A nurse-to-patient ratio of 1:4 applies for NICU/IMC patients requiring continuous cardiorespiratory monitoring, supplemental oxygen therapy, presence of a feeding tube, or the necessity of infusions or phototherapy. The treatment, nursing care, and cleaning services were provided by assigned and specialized teams.

### Definitions and Data acquisition

A patient hospital stay (PHS) was defined as continuous inpatient episode on the NICU/IMC. Leaving the unit (discharge or transfer to another ward/hospital) ended the PHS; any return to the NICU/IMC started a new PHS. Moves within the NICU/IMC complex did not create a new PHS. *Klebsiella pneumoniae* (Kp) prevalence was defined as the number of Kp patients per 100 patients, incidence density as Kp patient hospital stays (PHSs) per 1.000 patient days. A Kp case was any PHS with at least one Kp-positive specimen. Nosocomial Kp carriage was defined as ≥48 hours of NICU/IMC stay. Community-acquired Kp PHSs included acquisition in an outpatient or non-nosocomial setting (i.e. birth-related, in another ward of the same hospital, or an external hospital). Any additional PHSs of patients who already had a nosocomial PHS were generally not counted as new PHSs, as the endpoint had already been reached. One exemption was made for a patient with clear acquisition of a separate Kp strain during an additional PHS, indicated by an ESBL phenotype absent in the earlier PHS isolate; this was counted as a distinct PHS. The *time at risk* for Kp acquisition was defined as the number of days from admission to first detection of Kp in any microbiologic sample. If divergent Kp genotypes were identified in a patient during one NICU stay, the *time at risk* was defined from admission until detection of the first positive sample. The detection of a new Kp genotype in a Kp positive patient who returned to the NICU for a second stay, was considered as a new Kp hospital stay. A Kp infection was defined as the presence of Kp in a diagnostic microbiological specimen, such as a blood culture, combined with at least five days of antibiotic treatment, and clinical documentation. The microbiological laboratory prospectively documents every new Kp detection per PHS in samples from the NICU/IMC complex. Two authors independently screened this documentation to identify Kp positive specimens and corresponding patients and their patient hospital stays (PHSs). Microbiological metadata, such as antimicrobial resistance testing results, were retrieved directly from the database of the laboratory information system (m/LAB, Dorner, Müllheim, Germany). A third author extracted demographic and clinical data from patient records and the patient data management system until discharge. NICU patient profiles (nurse-to-patient ratio relative to birth weight) were obtained from primary care data sources. Hannover Medical School Information Technology provided overall patient numbers, PHSs, and patient days.

### IPC concept and surveillance

Following initial colonization screening upon admission, weekly microbiological screening was performed of respiratory secretions (or tracheal aspirates when applicable during invasive ventilation) and rectal swabs targeting Methicillin-resistant *Staphylococcus aureus* (MRSA), Vancomycin-resistant enterococci (VRE) and susceptible as well as resistant Gram-negative bacteria (e.g., Enterobacterales including Kp). All intensive care patients and those with an actual weight of <1500g in IMC were included. Diagnostic samples were taken based on clinical judgement. Upon Kp detection, a strict hygiene concept was followed, including use of personal protective equipment (gowns, gloves) during direct patient contact. In addition, patient isolation or cohorting was strongly recommended. Isolation was defined as single-room assignment and dedicated equipment. Cohorting measures were applied, when several patients tested Kp-positive, regardless of genotypes, as that information was available with delay. If a patient tested positive in the string test, the infection prevention and control (IPC) protocol recommended strict isolation in a single room. After discharge of a Kp positive patient, the room and reusable medical equipment were thoroughly disinfected, the non-reusable equipment was discarded.

Over the course of the study, the infection control team initiated regular IPC audits with the NICU staff and hygiene training and monitoring, such as hand hygiene compliance observations. Parents and visitors received repeated hygiene instructions. If clusters or transmissions were identified through the prospective epidemiological surveillance or genomic analysis, this triggered immediate IPC audits, on-site training, and sharing of genomic data, sample sites, and timelines with the NICU staff. Thereby, each Kp cluster prompted interventions, like enhanced training, compliance monitoring, and screening to improve awareness and control.

### Microbiological diagnostics

Diagnostics were conducted at the ISO 15189-accredited microbiology laboratory of Hannover Medical School. Screening for Gram-negative bacteria, including Kp, used MacConkey agar. For children over 12 weeks, additional MacConkey agar plates supplemented with either ceftazidime or cefotaxime were used to identify Gram-negative bacteria exhibiting an ESBL phenotype. Clinical samples were processed using both liquid and solid culture media according to standard procedures. Species identification was performed via matrix-assisted laser desorption/ionization time-of-flight mass spectrometry (Vitek MS system), and antimicrobial susceptibility testing by Vitek 2 system (both bioMérieux, Marcy-l'Étoile, France).
Kp isolates were tested with the string test to identify hypermucoviscous phenotypes (cut-off value: 5mm)[2]. Tellurite susceptibility was assessed via disk agar-diffusion test (Mueller-Hinton agar, disk with 10µl 3.5% tellurite solution, McFarland 0.5 bacteria solution, 18h incubation time, 36° incubation temperature) with laboratory-developed inhibition zone criteria (susceptible: ≥ 40mm; resistant ≤26mm; indeterminate >26-<40mm).

### Molecular analysis

Kp isolates were collected and re-cultured for molecular typing following ISO 15189 accreditation standards. Bacterial DNA was extracted (DNeasy PowerSoil Pro Kit, Qiagen, Venlo, Netherlands), libraries were prepared (Illumina DNA Prep, Illumina, San Diego, United States) and WGS was performed on an Illumina MiSeq platform (paired-end reads, 2 x 250 bp, MiSeq Reagent Kit v2, Illumina, San Diego, United States).
Initial de-novo assembly, quality control, multi-locus sequence typing (MLST), and core-genome MLST (cgMLST) were performed using Ridom SeqSphere+ (v10.0.4; Ridom, Münster, Germany) using the *K. pneumoniae sensu lato* cgMLST scheme (https://www.cgmlst.org/ncs/schema/Kpneumoniae4765/) with a cluster threshold of 15 allelic differences[3,4]. Isolates that failed workflow-defined quality thresholds were resequenced. Additional molecular analyses were conducted using Kleborate (v3.1.3)[5] via Pathogenwatch (v23.2.0; https://pathogen.watch).
For shared k-mer analysis (SKA) of publicly available genomes, metadata for all Kp genomes were downloaded from the Bacterial and Viral Bioinformatics Resource Center (BV-BRC) FTP server (ftp://ftp.bvbrc.org/genomes; accessed January 8, 2025), including information on isolation source, geographic origin and host. Metadata were filtered for MLST types identified in the present dataset, and corresponding assemblies were downloaded. Cohort and public assemblies were grouped by sequence type (ST), and for each ST, an input_sequences.txt file was generated. Using ska2 (v0.3.2)[6], we performed ska build and ska distance (with the --filter-ambiguous option) to compute pairwise single nucleotide polymorphism (SNP) distances within each ST. Distances <500 SNPs were retained and merged with the original BV-BRC metadata for downstream phylogenetic analyses. We used a cutoff of 20 SNPs defined by ska2 for cluster delineation.
Visualization of microbiological and molecular analyses were performed using R (v4.5.0) with ComplexHeatmap package for visualization[7].

### Machine learning

Reporting of machine learning models follows the TRIPOD+AI statement[8]. The outcome was Kp molecular cluster prevalence, calculated as the number of patients present on the ward in a specific calendar week carrying genomically clustered isolates.
Climate data were retrieved daily via the OpenWeatherMap One Call 3.0 API (https://openweathermap.org/api/one-call-3 ). Weekly aggregation was performed using the minimum and maximum values per week for minimum and maximum ambient daily temperatures, and the arithmetic mean for humidity, atmospheric pressure, and precipitation. Patient profile data were retrieved from shift-level documentation, as required by the German statutory health insurance (German Federal Joint Committee [G-BA]). First, the data were aggregated to daily medians, and subsequently to weekly values by taking the maximum daily median within each category. Patient categories were defined by the patients' birth weight, and the required nurse-to-patient ratio expressed the care level. For instance, a VLBW nurse-to-patient ratio of 1:2 referred to shifts in which one nurse was responsible for two VLBW patients.
Exploratory data analyses (EDA) were performed using Python and Altair visualization library (v5.5.0). To investigate delayed effects, we considered lags for climate and ward-load variables using visual EDA (weekly heatmaps; **Additional Figure 1**). We applied candidate two- and four-week lags by shifting the timestamps of the data forward by 14 or 28 days prior to aggregation. Data points with missing values introduced by this were removed from analysis. We fit candidate-lag models for two- and for weeks and compared hold-out $R^2$/RMSE. A 4-week lag for minimum temperature and ward-load metrics showed the strongest and most consistent association with cluster prevalence and the best hold-out fit (data for two-week candidate-lag models not shown).

The machine-learning models were applied primarily as statistical methods to explore and quantify associations between external drivers and Kp molecular cluster prevalence, explicitly not intended or validated as tools for real-world prediction or clinical forecasting. A season-aware train-test partition was created by random splitting (random seed=42) within each meteorological season (Winter, Spring, Summer, Autumn), resulting in a training set (80%) and a hold-out test set (20%). Predictive modelling employed penalized linear regression models (Ridge, Lasso, ElasticNet), random forest, and extreme gradient boosting (XGBoost). Random forest models and one XGBoost model included 24 features. Climate-related variables comprised humidity, precipitation, minimum daily outdoor temperature (temp_min), and maximum daily outdoor temperature (temp_max), each entered as contemporaneous and 4-week lagged values. Patient-care–related variables included the weekly peak numbers of very low birth weight (VLBW, <1500 g) infants and infants with birth weight >1500 g (HBW), as well as their distribution across nurse-to-patient ratios of 1:1, 1:2, and 1:4. For all patient-care features, both contemporaneous and 4-week lagged values were included. A reduced XGBoost model included only the six most influential features determined by initial training data screening, while feature reduction was guided by domain knowledge and iterative model comparison within the observed time frame (four-week lagged minimum temperature, four-week lagged maximum temperature, four-week lagged number of VLBW infants with nurse-to-patient-ratio of 1:2, four-week lagged number of patients with birth weight >1500g (HBW), current number of VLBW patients, and current humidity). All predictors used for a specific week were available at or before that week; no future information was used. Models were implemented in Python using scikit-learn (v1.7.0)[9] and XGBoost (v3.0.2)[10]. Model hyperparameters are detailed in **Additional Table 1**.

Model performance was evaluated using the coefficient of determination ($R^2$), root mean squared error (RMSE), and mean absolute error (MAE). Performance metrics were calculated exclusively on the hold-out dataset, and 95% confidence intervals (CI) were generated by bootstrapping (800 replicates, seed=42) without model refitting, therefore reflecting uncertainty of the performance estimate within the fixed trained model rather than variability due to model refitting.

Model interpretation was performed using Shapley Additive Explanations (SHAP; v0.48.0)[11]. Global feature importance was visualized via SHAP beeswarm plots on the hold-out data. To investigate the relative importance of each predictor across seasons, absolute SHAP values were computed and averaged separately within each season and then visualized as a heat map. Learning curves were created to characterize model generalizability by retraining the models using incrementally increasing fractions of the training data with season-stratified resampling, visualizing median performance metrics and interquartile ranges.

**Additional Table 1. Hold-out performances and hyperparameters of trained machine learning models.**

| Model | ElasticNet | Lasso | Ridge | RandomForest | XGBoost | XGBoost_Top6 |
|---|---|---|---|---|---|---|
| Features | 24 | 24 | 24 | 24 | 24 | 6 |
| R2 | -0.178 (-0.620 – 0.008) | -0.178 (-0.620 – 0.008) | 0.435 (0.197 – 0.579) | 0.784 (0.637 – 0.857) | 0.768 (0.644 – 0.836) | 0.801 (0.697 – 0.861) |
| RMSE | 1.913 (1.379 – 2.390) | 1.913 (1.379 – 2.390) | 1.324 (0.994 – 1.628) | 0.820 (0.581 – 1.046) | 0.849 (0.639 – 1.032) | 0.786 (0.581 – 0.976) |
| MAE | 1.481 (1.023 – 1.951) | 1.481 (1.023 – 1.951) | 1.085 (0.825 – 1.395) | 0.631 (0.448 – 0.844) | 0.665 (0.464 – 0.864) | 0.610 (0.435 – 0.793) |
| Hyper-parameters | { "alpha":2.7, "l1_ratio":0.4, "max_iter":5000 } | { "alpha":1.3, "max_iter":5000 } | { "alpha":100 } | { "max_depth":12, "max_features":0.4, "min_samples_leaf":1, "n_estimators":1000, "random_state":42 } | { "colsample_bytree":0.8, "learning_rate":0.01, "max_depth":2, "random_state":42, "reg_alpha":5, "reg_lambda":10, "subsample":0.8 } | { "colsample_bytree":0.8, "learning_rate":0.01, "max_depth":2, "random_state":42, "reg_alpha":5, "reg_lambda":10, "subsample":0.8 } |

Hold-out performance (20% season-stratified split, seed = 42) expressed as point estimate ± 95% bootstrap CI (800 resamples). Hyperparameter column lists all non-default settings of each trained model.

**Additional Table 2. Epidemiological and clinical parameters during time at risk of PHSs with nosocomial and community-acquired Kp.**

| Parameter | PHSs with nosocomial Kp (A) | PHSs with community-acquired Kp (B) | *P*-value[b] (A vs. B) |
|---|---|---|---|
| **Epidemiological parameters** | **n=58** | **n=24** | |
| Total PHSs, n ([a]) | 58 (57) | 24 (24) | - |
| Length of PHSs, median [IQR], days | 32.8 [17.1 – 62.4] | 6.15 [2.0 – 26.7] | **<.0001** |
| time at risk of PHSs, median [IQR], days | 12.5 [6.75 – 25.5] | 1.0 [0 – 2.0] | **<.0001** |
| **Patient associated parameters** | **n=57** | **n=24** | |
| VLBW, n (%) | 38 (66.7) | 7 (29.2) | **.002** |
| GA, median [IQR], days (weeks) | 208 (29.7) [188 (26.9) – 237 (33.9)] | 245 (35.0) [207.5 (29.6) – 272.8 (39.0)] | **.005** |
| Birth weight, median [IQR], grams | 1300 [955 – 1878] | 2145 [1484 – 2948] | **.01** |
| Sex, Male, n (%) | 32 (56.1) | 12 (50.0) | .61 |
| Death, n (%) | 6 (10.5) | 0 (0) | .10 |
| **Clinical parameters during *time at risk*** | **n=58** | **n=24** | |
| Central venous catheter, n (%) | 25 (43.1%) | 9 (37.5%) | .63 |
| Peripheral venous catheter, n (%) | 56 (96.6%) | 18 (75.0%) | **.003** |
| Invasive ventilation, n (%) | 23 (39.7%) | 6 (25.0%) | .21 |
| Non-invasive ventilation, n (%) | 44 (75.9%) | 6 (25.0%) | **<.0001** |
| Transurethral catheter, n (%) | 10 (17.2%) | 1 (4.2%) | .1 |
| Surgery, n (%) | 8 (13.8%) | 0 (0%) | .06 |
| Systemic (intravenous) antibiotic therapy, n (%) | 25 (43.1%) | 4 (16.7%) | **.02** |
| Cefotaxime, n (%) | 11 (19.0%) | 0 (0%) | **.02** |
| Vancomycin, n (%) | 8 (13.8%) | 1 (4.2%) | .20 |
| Meropenem, n (%) | 9 (15.5%) | 1 (4.2%) | .15 |
| Tobramycin, n (%) | 22 (37.9%) | 3 (12.5%) | **.02** |

Abbreviations and definitions: PHS, patient hospital stay. Time at risk for Kp acquisition was defined as the number of days from admission to first detection of Kp in any microbiologic sample. Community-acquired acquisition comprised outpatient, birth-related or external nosocomial (i.e. acquired at another ward of the same hospital or at an external hospital) settings.
[a] Corresponding number of single patients. [b] Chi-square test for categorical parameters and Wilcoxon rank sum test for continuous parameters, respectively. Level of significance *P*<.05.

### Additional Table 3: Duration of clinical parameters in nosocomial Kp PHSs stratified by molecular cluster assignment.

| **Duration of clinical parameter** median [IQR], (days) | **Nosocomial Kp PHSs molecular cluster assignment (A)** | **Nosocomial Kp PHSs without molecular cluster assignment (B)** | ***P*-value[b] (A vs. B)** |
|---|---|---|---|
| Total PHSs, n ([a]) | 37 (37) | 21 (21) | **-** |
| Central venous catheter | 0 [0 – 12.5] | 0 [0 – 11.5] | .87 |
| Peripheral venous catheter | 7.0 [6.0 – 10.5] | 5.0 [5.0 – 8.0] | **.01** |
| Invasive ventilation | 0 [0 – 6.5] | 0 [0 – 4.0] | .69 |
| Noninvasive ventilation | 8.0 [4.5 – 22.0] | 2.0 [0 – 7.0] | **.01** |
| Transurethral catheter | 0 [0 – 0] | 0 [0 – 2.0] | .14 |
| Cefotaxime | 0 [0 – 0] | 0 [0 – 0] | .48 |
| Vancomycin | 0 [0 – 0] | 0 [0 – 1.0] | .18 |
| Meropenem | 0 [0 – 0] | 0 [0 – 0] | .82 |
| Tobramycin | 0 [0 – 4.0] | 0 [0 – 0.5] | .05 |

Abbreviations: PHS, patient hospital stay.
[a] Corresponding number of single patients. [b] Wilcoxon rank sum test for continuous parameters, respectively. Level of significance ***P*<.05**.

### Additional Table 4. Isolate-level pheno-genomic metadata, patient/PHS linkage, and Pathogenwatch/Kleborate outputs.

(Separate Excel Data File "Additional File 2")

This table links each Kp isolate to the corresponding patient and patient-hospital-stay (PHS) and summarizes isolate metadata, genomic relatedness, phenotypes, and automated annotations from Pathogenwatch/Kleborate. The "PATHOGENWATCH Genome ID" column provides the public accession in Pathogenwatch.
Abbreviations: PHS, patient-hospital-stay; MLST, multilocus sequence typing; cgMLST, core-genome MLST; SKA2_cutoff20, SKA clustering at ≤20 SNPs; EUCAST, European Committee on Antimicrobial Susceptibility Testing; SAM ampicillin/sulbactam; TZP piperacillin/tazobactam; CXM cefuroxime; CTX cefotaxime; CAZ ceftazidime; MEM meropenem; GEN gentamicin; CIP ciprofloxacin; SXT trimethoprim/sulfamethoxazole; String, hypermucoviscosity string test; Tellurite, disk diffusion tellurite susceptibility; YbST/CbST/AbST/SmST/RmST, Kleborate sequence types for yersiniabactin/colibactin/aerobactin/salmochelin/rmpADC; K-locus/O-locus, capsule and O-antigen loci.

### Additional Table 5. SKA2 SNP distances between study *Klebsiella pneumoniae* sensu stricto genomes and BV-BRC public genomes, with accessions and isolation metadata.

(Separate Excel Data File "Additional File 2")

This table lists, for each *Klebsiella pneumoniae* sensu stricto genome from this study, the pairwise single-nucleotide polymorphism (SNP) distance to publicly available genomes from the Bacterial and Viral Bioinformatics Resource Center (BV-BRC), computed with SKA2. Distances are reported as SNP counts; smaller values indicate closer genetic relatedness. For each BV-BRC genome, accession identifiers and isolation metadata are provided as supplied by BV-BRC. Study isolate accessions correspond to the Pathogenwatch Genome IDs listed in Additional Table 4. Missing or unavailable metadata are shown as NA.

### Additional Figure 1. Weekly cluster prevalence and concurrent climate and patient-care load.

Top: Weekly number of patients with *K. pneumoniae complex* isolates assigned to a molecular cluster (bars), based on cgMLST thresholds. Colors match Figure 1; weeks without *K. pneumoniae complex* detection are left blank.
Bottom: Heatmaps showing weekly summary statistics for climate and patient-care parameters, aligned on the same calendar week (x-axis) × year (y-axis) grid. Meteorological data were aggregated from daily OpenWeatherMap calls using the minimum of daily minima for weekly minimum temperature (°C), the maximum of daily maxima for weekly maximum temperature (°C), and the arithmetic mean for weekly humidity (%), pressure (hPa), and precipitation (mm). Patient-care variables include ward census and peak nurse-to-patient loads. VLBW = very low birth weight (<1500 g); HBW = birth weight >1500 g ("higher birth weight"). Labels such as "VLBW :1" and "HBW :4" denote maximum weekly counts at nurse-to-patient ratios of 1:1, 1:2, or 1:4. Color scales are independent per heatmap.

### Additional Figure 2. SHAP dependence plots of key features in the reduced-feature XGBoost model.

SHAP dependence plots for selected features, showing non-linear relationships with model predictions of molecular cluster prevalence. Each point represents a weekly observation. X-axis: feature value; Y-axis: SHAP value (feature contribution). Color scale reflects feature intensity. Features include: [Tmin (lagged)] minimum temperature, 4-week lag; [Tmax (lagged)] maximum temperature, 4-week lag; [RH] current humidity; [HBW (lagged)] peak number of patients with birth weight >1500g ("higher birth weight") treated, 4-week lag; [VLBW :2 (lagged)] Very-low-birth-weight (<1500g) patients treated with a 1:2 nurse-to-patient ratio (1 nurse per 2 patients), 4-week lag; [VLBW] current peak number of very-low-birth-weight patients treated (all nursing ratios). Grey bars are histograms indicating the distribution of the respective feature value.

### Additional Figure 3. Learning curves for XGBoost (Top-6) and random forest (all 24 features).

Learning curves were generated using GroupShuffleSplit cross-validation (10 splits; 70% training/30% test per split; grouping by season). For each training-set fraction (0.1–1.0), models were refit within each split. Lines show the median $R^2$ across splits for training (black) and cross-validation/test (gray); shaded bands indicate ± half of the interquartile range across splits.

### Additional References.


1. Rossi R, Bauer NH, Becke-Jakob K, et al. Empfehlungen für die strukturellen Voraussetzungen der perinatologischen Versorgung in Deutschland (Entwicklungsstufe S2k, AWMF-Leitlinien-Register Nr. 087–001, März 2021). *Z Geburtshilfe Neonatol.* 2021;225(4):306-319.

2. Tan TY, Cheng Y, Ong M, Ng LS. Performance characteristics and clinical predictive value of the string test for detection of hepato-virulent Klebsiella pneumoniae isolated from blood cultures. *Diagn Microbiol Infect Dis.* 2014;78(2):127-128.

3. Weber RE, Pietsch M, Frühauf A, et al. IS26-Mediated Transfer of bla (NDM-1) as the Main Route of Resistance Transmission During a Polyclonal, Multispecies Outbreak in a German Hospital. *Front Microbiol.* 2019;10:2817.

4. Jünemann S, Sedlazeck FJ, Prior K, et al. Updating benchtop sequencing performance comparison. *Nat Biotechnol.* 2013;31(4):294-296.

5. Lam MMC, Wick RR, Watts SC, Cerdeira LT, Wyres KL, Holt KE. A genomic surveillance framework and genotyping tool for Klebsiella pneumoniae and its related species complex. *Nat Commun.* 2021;12(1):4188.

6. Derelle R, von Wachsmann J, Mäklin T, et al. Seamless, rapid, and accurate analyses of outbreak genomic data using split k-mer analysis. *Genome Res.* 2024;34(10):1661-1673.

7. Gu Z, Eils R, Schlesner M. Complex heatmaps reveal patterns and correlations in multidimensional genomic data. *Bioinformatics.* 2016;32(18):2847-2849.

8. Collins GS, Moons KGM, Dhiman P, et al. TRIPOD+AI statement: updated guidance for reporting clinical prediction models that use regression or machine learning methods. *BMJ.* 2024;385:e078378.

9. Pedregosa F, Varoquaux G, Gramfort A, et al. Scikit-learn: Machine Learning in Python. *Journal of Machine Learning Research.* 2011;12:2825-2830.

10. Chen T, Guestrin C. XGBoost: A Scalable Tree Boosting System. Proceedings of the 22nd ACM SIGKDD International Conference on Knowledge Discovery and Data Mining; 13. August 2016, 2016.

11. Lundberg SM, Lee SI. A unified approach to interpreting model predictions. *Advances in neural information processing systems.* 2017;30.

Additional Figure 1

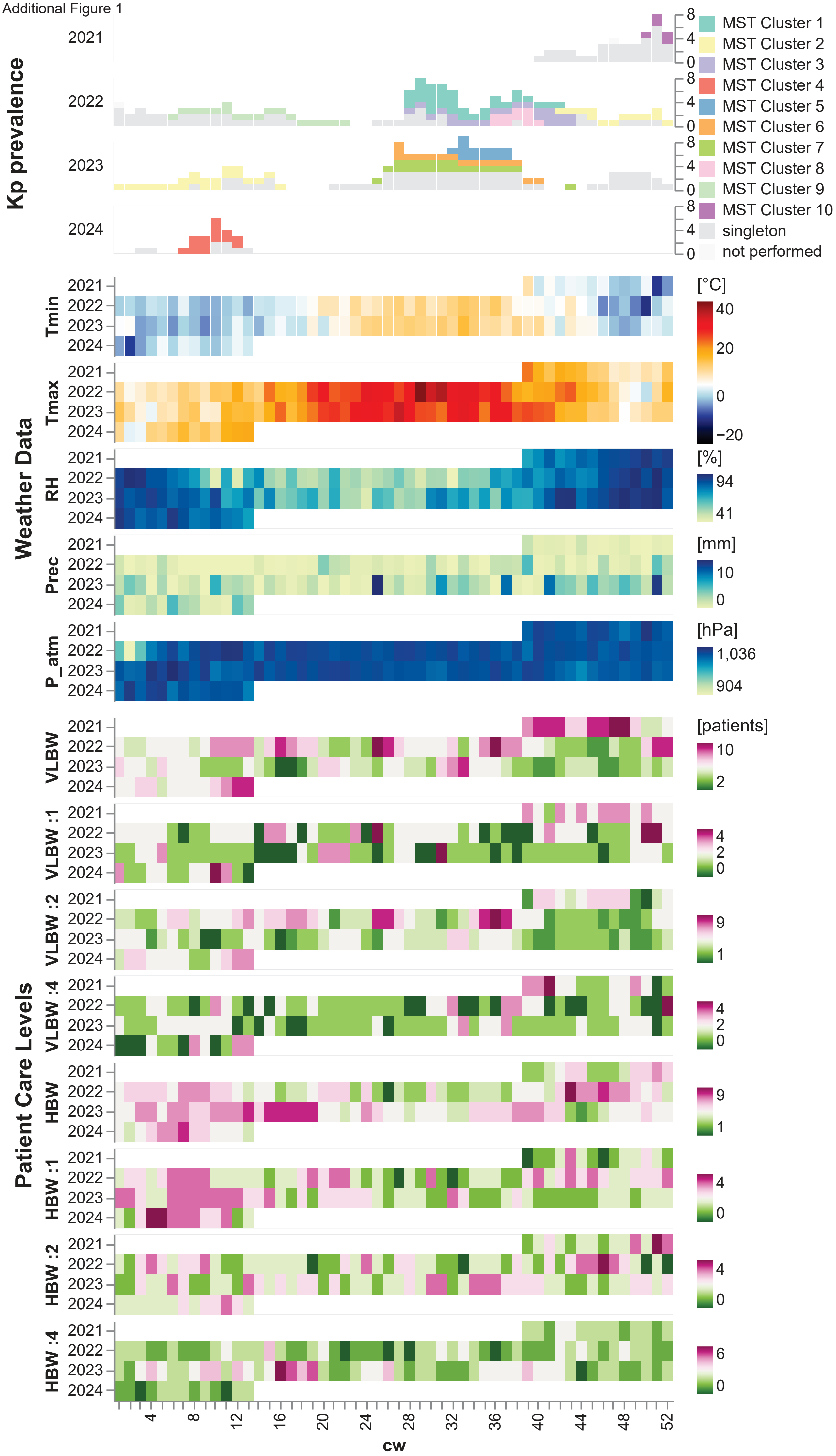

Additional Figure 2

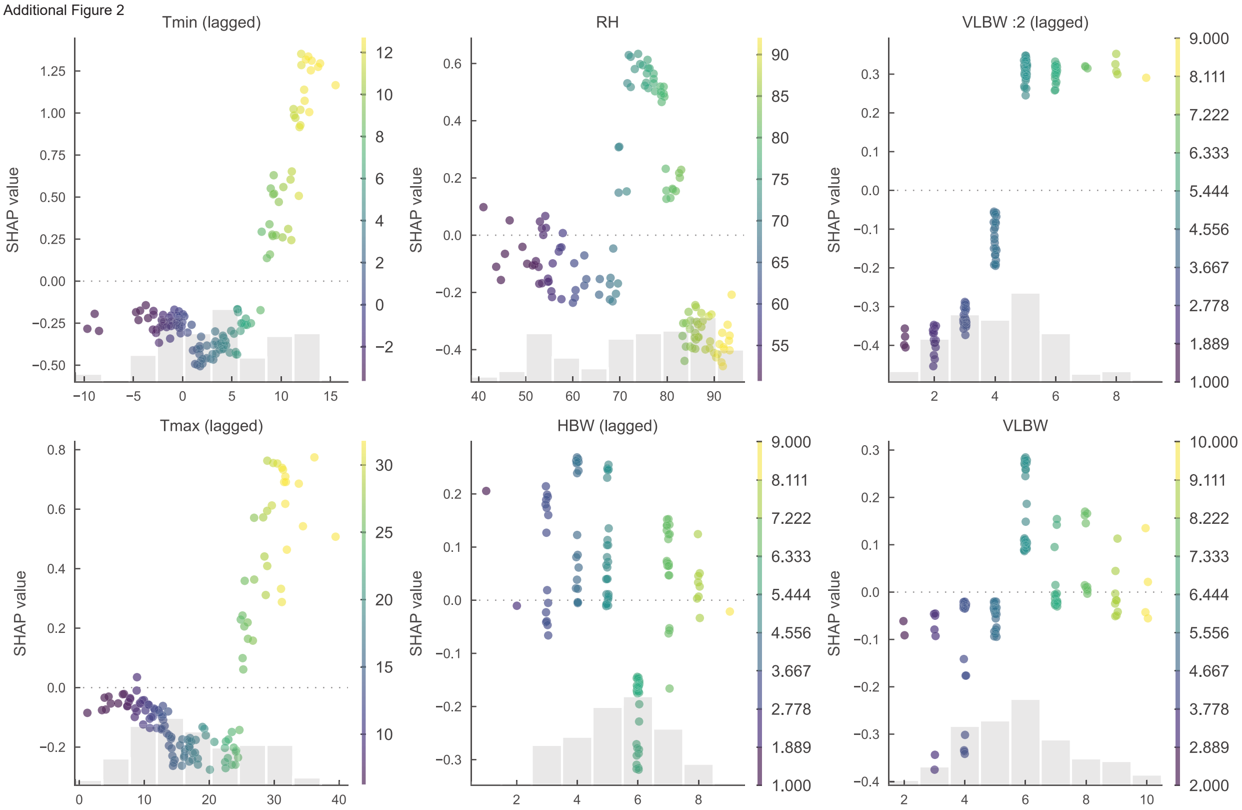

Additional Figure 3

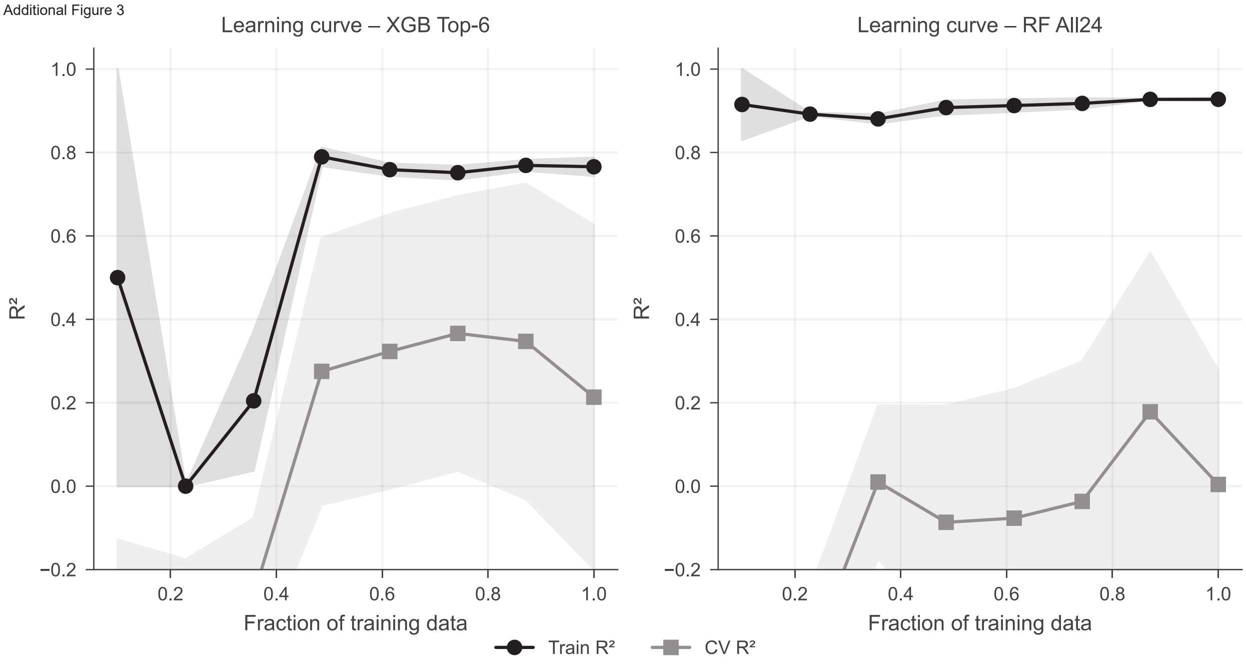
